# Metabolic Disease and The Risk of Post-COVID Conditions: A Retrospective Cohort Study

**DOI:** 10.1101/2024.03.26.24304845

**Authors:** Wubin Xie, Heather E. Hsu, Paul R. Shafer, Meghan I. Podolsky, Andrew C. Stokes

**Author notes:** **Address correspondence to:** Andrew C. Stokes, 801 Massachusetts Ave., Crosstown 362, Boston, MA 02118.

## Abstract

**Objective:** To examine the influence of having a baseline metabolic disorder (diabetes, hypertension, and/or obesity) on the risk of developing new clinical sequelae potentially related to SARS-CoV-2 in a large sample of commercially insured adults in the US.

**Design, setting, and participants:** Deidentified data were collected from the IBM/Watson MarketScan Commercial Claims and Encounters (CCAE) Databases and Medicare Supplemental and Coordination of Benefits (MDCR) Databases from 2019 to 2021. A total of 839,344 adults aged 18 and above with continuous enrollment in the health plan were included in the analyses. Participants were grouped into four categories based on their COVID-19 diagnosis and whether they had at least one of the three common metabolic disorders at baseline (diabetes, obesity, or hypertension).

**Measures and methods:** ICD-10-CM codes were used to determine new symptoms and conditions after the acute phase of SARS-CoV-2 infection, defined as ending 21 days after initial diagnosis date, or index period for those who did not have a COVID-19 diagnosis. Propensity score matching (PSM) was used to create comparable reference groups. Cox proportional hazard models were conducted to estimate hazard ratios and 95% confidence intervals.

**Results:** Among the 772,377 individuals included in the analyses, 36,742 (4.8%) without and 20,912 (2.7%) with a baseline metabolic disorder were diagnosed with COVID-19. On average, COVID-19 patients with baseline metabolic disorders had more 2.4 more baseline comorbidities compared to those without baseline metabolic disorders. Compared to adults with no baseline metabolic condition, the risks of developing new clinical sequelae were highest among COVID-19 patients with a baseline metabolic condition (HRs ranging from 1.51 to 3.33), followed by those who had a baseline metabolic condition but with no COVID-19 infection (HRs ranging from 1.33 to 2.35), and those who had COVID-19 but no baseline metabolic condition (HRs ranging from 1.34 to 2.85).

**Conclusions:** In a large national cohort of commercially insured adults, COVID-19 patients with a baseline metabolic condition had the highest risk of developing new clinical sequelae post-acute infection phase, followed by those who had baseline metabolic condition but no COVID-19 infection and those who had COVID-19 but no baseline metabolic disorder.

## Introduction

Since its first identification at the end of 2019, severe acute respiratory syndrome coronavirus 2 (SARS-CoV-2) has infected over 676 million people globally.^1^ An estimated 6.8 million people have died from coronavirus disease 2019 (COVID-19), which is caused by the SARS-CoV-2 virus. The US has recorded the largest number of deaths and confirmed COVID-19 cases worldwide. By March 10, 2023, there were over 103 million confirmed cases and 1.1 million deaths in the US.^1^ After the acute phase of infection, a subset of people with COVID-19 experience post-acute sequelae of COVID-19 (PASC), adding to the disease burden. While estimations vary widely, a recent systematic review reported that 54% of COVID-19 survivors experienced PASC up to 6 months after recovery.^2^

Obesity is highly prevalent in most countries^3^ including the US, where over 40% of the population is affected. Over 120 million adults have either prediabetes or diabetes, with many undiagnosed, and nearly half of US adults have hypertension.^4,5^ Underlying metabolic conditions are common in patients with COVID-19.^6–9^ Patients with obesity and impaired metabolic health are susceptible to developing severe symptoms of COVID-19,^8,10^ and COVID-19 infection could worsen existing metabolic disorders or lead to new metabolic diseases.^11^ Some studies with relatively small sample sizes have reported a higher risk of PASC in those with obesity^12^ and diabetes.^13,14^ However, another study with a sample of older adults found no association between diabetes and PASC.^15^ Little is known about how baseline metabolic disorders interact with SARS-CoV-2 infection to modify risk of developing new clinical sequelae.

The present study examined the influence of baseline metabolic disorders on the risk of developing new clinical sequelae potentially related to SARS-CoV-2 in a large sample of commercially insured adults.

## Methods

### Data source and cohort identification

Deidentified data were collected from the IBM/Watson MarketScan Commercial Claims and Encounters (CCAE) Databases, and Medicare Supplemental and Coordination of Benefits (MDCR) Databases for the from January 1, 2019 to July 31, 2021. These databases include inpatient, outpatient, and pharmacy claims, as well as enrollment data from large employers and health plans across the US who provide private healthcare coverage to employees, spouses, and dependents.

The study population included 772,377 adults aged 18 and above who were continuously enrolled from January 1, 2019, to the index date (**Supplementary Figure 1**). The index date for COVID-19 patients was defined as the date of first diagnosis of COVID-19 at an outpatient or an inpatient encounter, identified using ICD-10-CM diagnosis code U07.1. Individuals with a diagnosis code of B34.2 or B97.29 during January 1 and March 31, 2020 were included because many healthcare professionals used these codes before the Centers for Disease Control and Prevention recommended U07.1 as the primary code for clinical diagnosis on April 1, 2020.^16^ Individuals from the comparison group who did not have a recorded COVID-19 diagnosis were randomly assigned to an index date drawn from COVID-19 patients. The approach has been described in detail in a previous publication using another commercial claims dataset.^17^

### Assessment of baseline metabolic diseases

Using claims between January 1, 2019 and the index date, baseline metabolic diseases (diabetes, hypertension, and obesity) were determined by ICD-10-CM codes derived from the Elixhauser comorbidity index. Given that these conditions are often clustered, we created a binary exposure capturing whether an individual had at least one of these conditions. The ICD-10-CM codes used to define each of the metabolic conditions are available in **Supplementary Table 1**. We then created four joint exposure patterns based on COVID-19 infection and baseline metabolic disease. The groups were 1) “*none”*, no COVID-19 infection and no baseline metabolic disease; 2) “*COVID-19”*, COVID-19 infection and no baseline metabolic disease; 3) “*any metabolic condition”*, no COVID-19 infection and at least one baseline metabolic disease; and 4) “*both”*, COVID-19 infection with at least one baseline metabolic disease. Similar classifications were created for each of the three metabolic conditions separately.

### Outcomes

Informed by previous studies,^17,18^ we used ICD-10-CM codes to identify selected new clinical diagnoses from claims of inpatient and outpatient encounters. A clinical diagnosis was considered an incident case if the defining diagnostic codes recorded after index date and not before. Details are provided in **Supplementary Table 1**.

### Covariates

We used data from annual enrollment summaries to ascertain age (18-24, 25-34, 35-44, 45-54, 55-64, 65+ years), sex (male, female), region (Northeast, North Central, South, West), COVID-19 diagnosis period (January-March 2020, April-June 2020, July-September 2020, October-December 2020, January-March 2021, April-July 2021). Existing comorbidities were derived from the Charlson and Elixhauser indices, determined by inpatient and outpatient claims between January 1, 2019 and immediately before the index date. We used ELIXHAUSER and CHARLSON Stata modules to create binary indicators of each condition.^19,20^ These comorbidities included congestive heart failure, cardiac arrhythmias, valvular disease, pulmonary circulation disorders, peripheral vascular disorders, paralysis, other neurological disorders, chronic pulmonary disease, hypothyroidism, renal failure, liver disease, peptic ulcer disease, HIV/AIDS, lymphoma, metastatic cancer, solid tumor without metastasis, rheumatoid arthritis, coagulopathy, weight loss, fluid and electrolyte disorder, blood loss anemia, deficiency anemia, alcohol abuse, drug abuse, psychoses, depression, myocardial infraction, cerebrovascular disease, dementia, and hemiplegia/paraplegia.

### Statistical analysis

We calculated incidence rates for all new clinical sequelae. Individuals with existing diagnoses of interest before index date or those who developed these conditions during the acute phase of infection were excluded from the calculation. A series of models were performed to estimate the associations between COVID-19 infection, baseline metabolic conditions, and risk of developing clinical sequelae, using individuals who had no baseline metabolic disorders and no COVID-19 diagnosis as reference. We compared the relative risk of new clinical sequelae among COVID-19 patients with a baseline metabolic condition compared to the three reference groups. We assessed the interaction between baseline metabolic condition and COVID-19 severity, proxied by whether an inpatient or ICU admission was recorded. We then stratified the analyses by age (18 to 64 years and 65 years and above) and sex. Finally, we evaluated joint exposure to COVID-19 and each baseline metabolic condition separately.

Hazard ratios were calculated with 95% confidence intervals (CI) using Cox proportional hazards models. Individuals were followed up until the occurrence of outcome of interest (each being evaluated separately), loss of health insurance coverage, or end of study (July 31, 2021). The proportional hazards assumption was tested with Schoenfeld residuals.^21^ Models were adjusted for all covariates described above. Post-hoc tests were performed to compare differences in coefficients among exposure groups. All tests were 2-tailed and were adjusted for multiple comparisons using Holm’s sequential Bonferroni procedure,^22^ and statistical significance was set at p<0.05. Analyses were conducted in Stata 17 (StataCorp).

In a sensitivity analysis, we tested an alternative way to address confounding using PSM to create reference cohorts with matched baseline characteristics with all the covariates indicated above.^23^ PSM with 1:1 matching to nearest neighbor without replacement was performed. Standardized mean difference of <0.1 was regarded as well matched. Any unbalanced characteristics were included as additional covariates.

## Results

### Sample characteristics and metabolic conditions at baseline

Among the 772,377 individuals included in the analysis, 36,742 (4.8%) were diagnosed with COVID-19 and did not have any of the baseline metabolic disorders and 20,912 (2.7%) had both COVID-19 and at least one metabolic disorder at baseline. Most of the analytic sample (n=510,110) were free of COVID-19 and baseline metabolic disorders and 204,613 (26.5%) individuals had at least one baseline metabolic disorder but were not diagnosed with COVID-19. The mean age was 39.6 (standard deviation [SD] 14.1) for the COVID-19 only group and 52.8 (SD 14.1) for COVID-19 patients with a baseline metabolic condition. Across the groups, most individuals were from the South (44.5%-55.5%). The proportion of COVID-19 cases identified in the first quarters of 2021 were lower than the last quarter of 2020 due to incompleteness of 2021 data.

Individuals with COVID-19 and no baseline metabolic disorders had a similar number of comorbidities compared with those who had no COVID-19 or baseline metabolic disorders (0.3 versus 0.4). Individuals with a baseline metabolic disorder appeared to have more comorbidities (2.8 among those without COVID-19 and 3.1 among those with COVID-19) than those without a metabolic condition (0.3 among those without COVID-19 and 0.4 among those with COVID-19).

### Risk of post-acute sequelae of COVID-19 by baseline metabolic disorder

COVID-19 patients with a baseline metabolic condition had the highest incidence of post-acute clinical sequelae, followed by those who had a baseline metabolic condition but no COVID-19 infection, those who had COVID-19 but no baseline metabolic disorder, and those who had none (**Table 2**). For almost all outcomes, the crude incidence rates were higher for COVID-19 patients with baseline metabolic conditions compared with those without. The pattern of higher incidence associated with baseline metabolic conditions was also observed among the population without COVID-19. Cumulative incidence rates for selected outcomes are presented in **Figure 2**. Common outcomes with high incidence rates included joint pain (175.2 cases per 1,000 person-years in the dual-exposed group), dorsalgia (155.7 cases per 1,000 person-years), dyspnea (100.7 cases per 1,000 person-years), anxiety disorder (103.4 cases per 1,000 person-years), and chest pain (836 cases per 1,000 person-years).

**Figure 1.**
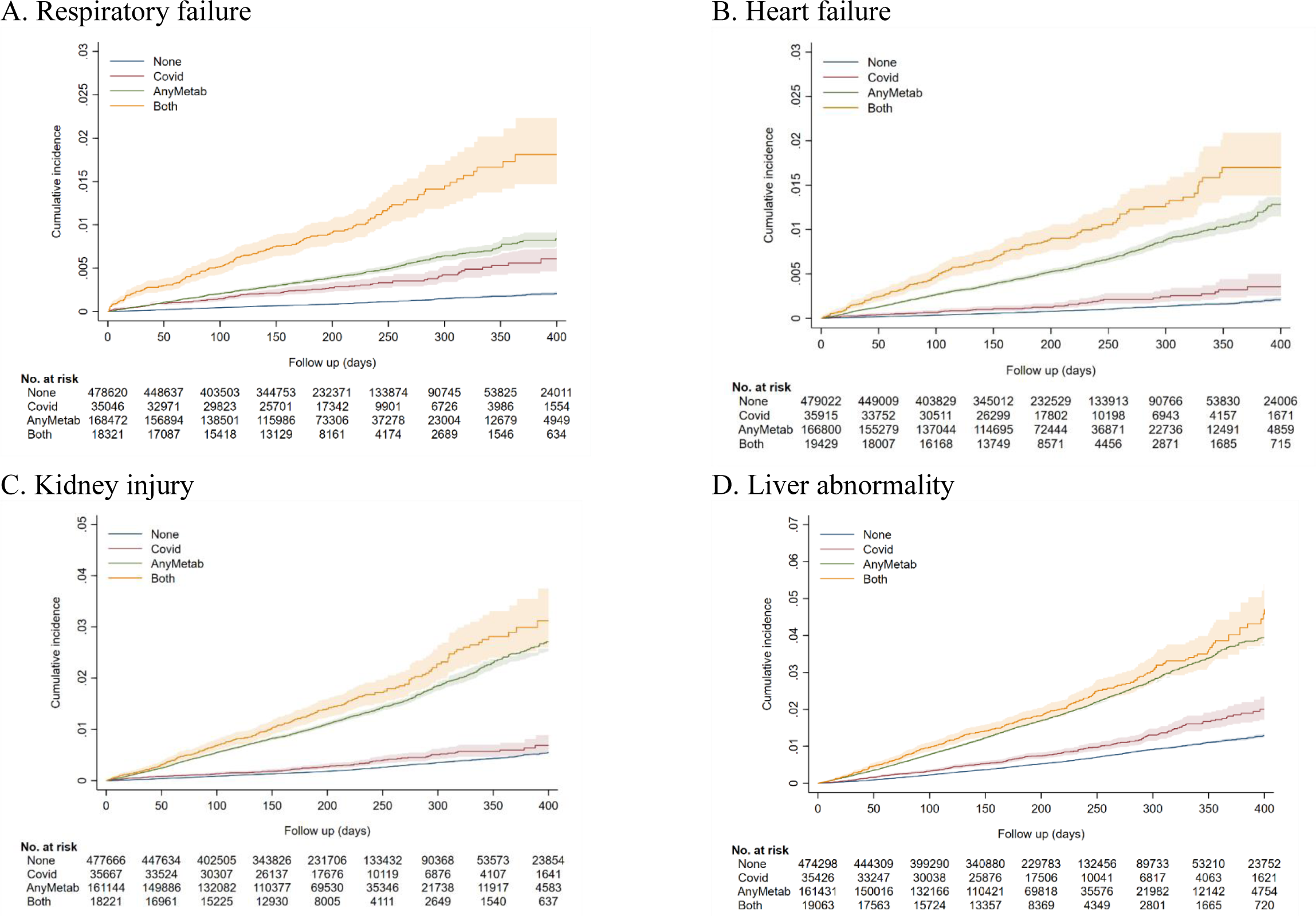
Cumulative incidence rates of selected post-acute sequelae of COVID-19 by baseline metabolic condition and COVID-19 infection. None: Neither COVID-19 diagnosis nor pre-existing diagnosis of obesity, diabetes, and/or hypertension. Covid: COVID-19 diagnosis, no pre-existing diagnosis of obesity, diabetes, and/or hypertension. AnyMetab: Diagnosis of obesity, diabetes, or hypertension with no COVID-19 diagnosis. Both: Both COVID-19 diagnosis and pre-existing diagnosis of obesity, diabetes, and/or hypertension.

**Figure 2.**
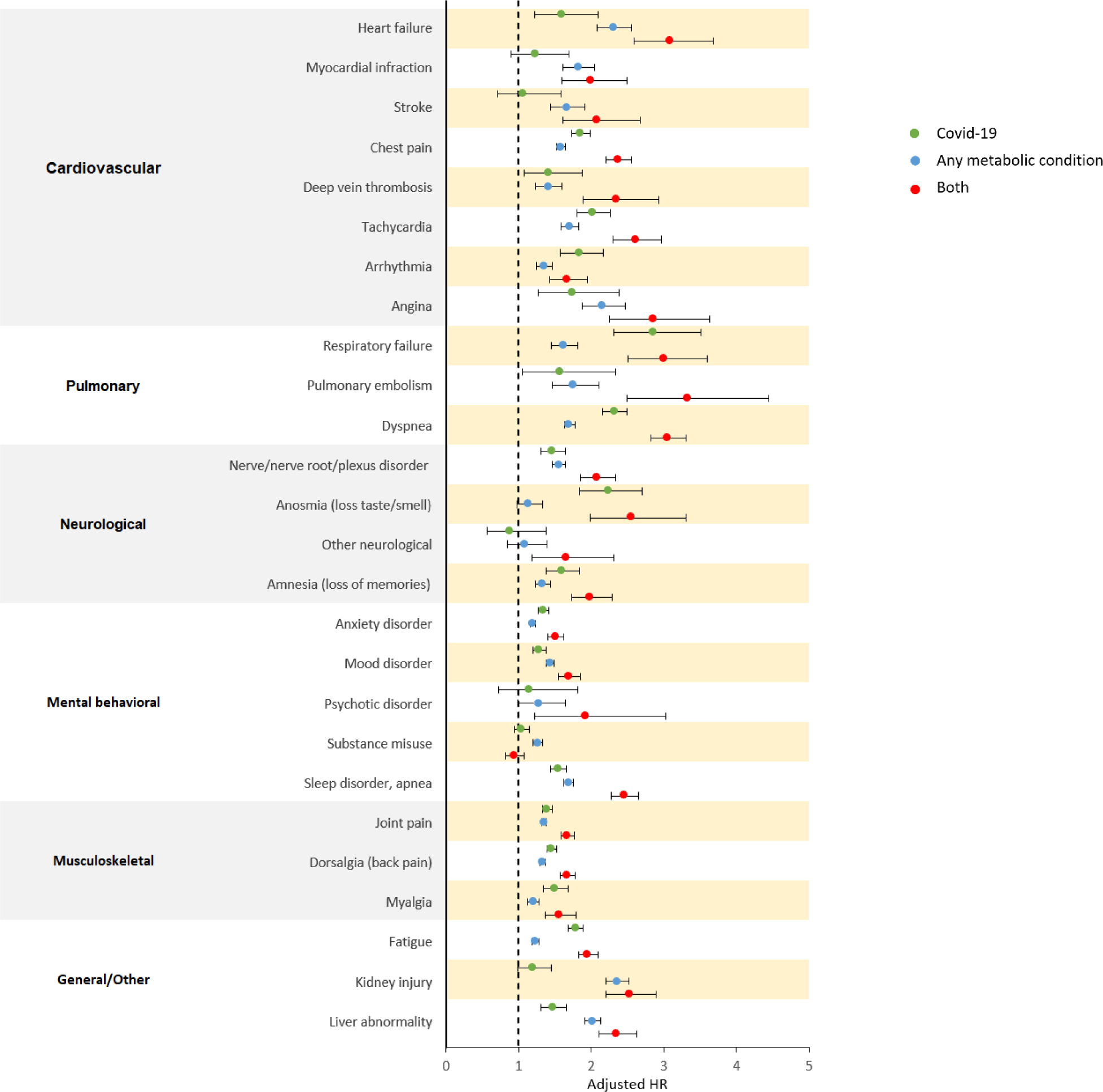
Multivariable adjusted hazard ratios for post-acute sequelae of COVID-19. Any metabolic condition included at least one of three baseline metabolic disorders (diabetes, hypertension, obesity) diagnosed between January 1, 2019 and index date. Mutually exclusive joint-exposure categories were based on baseline metabolic condition (yes/no) and COVID-19 diagnosis (yes/no), classifying participants as having neither (referent), COVID-19 only, baseline metabolic condition only, and both COVID-19 diagnosis and baseline metabolic condition. Models were adjusted for age (18-24, 25-34, 35-44, 45-54, 55-64, 65+), sex (male, female), region (northeast, north central, south, west), COVID-19 diagnosis time period (January-March 2020, April-June 2020, July-September 2020, October-December 2020, January-March 2021, April-July 2021), and comorbidities (congestive heart failure, cardiac arrhythmias, valvular disease, pulmonary circulation disorders, peripheral vascular disorders, paralysis, other neurological disorders, chronic pulmonary disease, hypothyroidism, renal failure, liver disease, peptic ulcer disease, HIV/AIDS, lymphoma, metastatic cancer, solid tumor without metastasis, rheumatoid arthritis, coagulopathy, weight loss, fluid and electrolyte disorder, blood loss anemia, deficiency anemia, alcohol abuse, drug abuse, psychoses, depression, myocardial infraction, cerebrovascular disease, dementia, hemiplegia/paraplegia).

**Table 1.** Unmatched sample characteristics by COVID-19 infection and baseline metabolic condition, IBM MarketScan 2019-2021.

| **Characteristics <sup>‡</sup>** | **None** | **COVID-19** | **Any Metabolic condition** | **Both** |
| --- | --- | --- | --- | --- |
| N | 510110 | 36742 | 204613 | 20912 |
| Age, mean (sd) | 41.3 (15.0) | 39.6 (14.1) | 55.3 (14.3) | 52.8 (14.1) |
| Age category |  |  |  |  |
| 18-24 | 96086 (18.8) | 7575 (20.6) | 4896 (2.4) | 550 (2.6) |
| 25-34 | 86974 (17.1) | 7001 (19.1) | 10899 (5.3) | 1470 (7.0) |
| 35-44 | 109753 (21.5) | 7907 (21.5) | 26719 (13.1) | 3332 (15.9) |
| 45-54 | 105641 (20.7) | 8117 (22.1) | 51051 (25.0) | 6066 (29.0) |
| 55-64 | 91518 (17.9) | 5459 (14.9) | 73482 (35.9) | 6932 (33.2) |
| 65+ | 19960 (3.9) | 677 (1.8) | 37478 (18.3) | 2556 (12.2) |
| Sex |  |  |  |  |
| Women | 245805 (48.2) | 16633 (45.3) | 98957 (48.4) | 9755 (46.6) |
| Men | 264305 (51.8) | 20109 (54.7) | 105656 (51.6) | 11157 (53.4) |
| Region |  |  |  |  |
| Northeast | 77452 (15.2) | 5847 (15.9) | 24461 (12.0) | 2539 (12.2) |
| North Central | 121748 (23.9) | 7741 (21.1) | 58964 (28.9) | 5063 (24.2) |
| South | 226361 (44.5) | 18631 (50.8) | 100744 (49.3) | 11591 (55.5) |
| West | 83465 (16.4) | 4472 (12.2) | 20085 (9.8) | 1697 (8.1) |
| Diagnosis |  |  |  |  |
| Jan - Mar, 2020 | 8230 (1.6) | 500 (1.4) | 1902 (0.9) | 240 (1.1) |
| Apr - Jun, 2020 | 55751 (10.9) | 3791 (10.3) | 14630 (7.2) | 1648 (7.9) |
| Jul - Sep, 2020 | 92263 (18.1) | 6529 (17.8) | 29514 (14.4) | 2899 (13.9) |
| Oct - Dec, 2020 | 195569 (38.3) | 13859 (37.7) | 84736 (41.4) | 8248 (39.4) |
| Jan - Mar, 2021 | 122793 (24.1) | 9190 (25.0) | 55904 (27.3) | 6083 (29.1) |
| Apr - Jul, 2021 | 35504 (7.0) | 2873 (7.8) | 17927 (8.8) | 1794 (8.6) |
| No. of comorbidity, mean (sd) | 0.3 (0.7) | 0.4 (0.8) | 2.8 (2.1) | 3.1 (2.4) |

SD: standard deviation. COVID-19=coronavirus disease 2019. Any metabolic condition consisted of a diagnosis of obesity, hypertension, and/or diabetes between January 1, 2019 and index date.
<sup>‡</sup>% and N presented unless indicated otherwise.

**Table 2.** Rates and hazard ratios for post-acute sequelae of COVID-19 by baseline metabolic condition and COVID-19 diagnosis.

| Outcomes | # of cases (Incidence per 1,000 person-years) |  |  |  | Adjusted Hazard Ratio (95% CI) |  |  |  |
| --- | --- | --- | --- | --- | --- | --- | --- | --- |
|  | None | COVID-19 | Any metabolic | Both | COVID-19 vs None | Any metabolic vs None | Both vs None |  |
| **Cardiovascular** |  |  |  |  |  |  |  |  |
| Heart failure | 625 (2.2) | 57 (2.8) | 1645 (15.8) | 170 (16.4) | 1.60 (1.22 - 2.10) | 2.30 (2.08 - 2.55) | 3.09 (2.59 - 3.68) | <sup>a,b</sup> |
| Myocardial infraction | 538 (1.9) | 41 (2.0) | 991 (9.3) | 99 (9.3) | 1.24 (0.90 - 1.70) | 1.82 (1.62 - 2.05) | 2.00 (1.59 - 2.50) |  |
| Stroke | 400 (1.4) | 26 (1.3) | 712 (6.6) | 79 (7.4) | 1.07 (0.72 - 1.59) | 1.66 (1.44 - 1.91) | 2.08 (1.61 - 2.67) | <sup>a</sup> |
| Chest pain | 7735 (27.7) | 968 (51.9) | 6045 (62.8) | 836 (95.2) | 1.85 (1.73 - 1.98) | 1.58 (1.52 - 1.64) | 2.37 (2.21 - 2.56) | <sup>a,b</sup> |
| Deep vein thrombosis | 587 (2.0) | 56 (2.7) | 649 (6.0) | 104 (9.7) | 1.42 (1.08 - 1.87) | 1.41 (1.24 - 1.60) | 2.35 (1.89 - 2.93) | <sup>a,b</sup> |
| Tachycardia | 2258 (7.9) | 334 (16.6) | 1907 (18.2) | 296 (28.9) | 2.02 (1.80 - 2.27) | 1.70 (1.58 - 1.83) | 2.61 (2.30 - 2.97) | <sup>a,b</sup> |
| Arrhythmia | 1534 (5.4) | 166 (8.1) | 1859 (18.4) | 191 (18.8) | 1.84 (1.57 - 2.16) | 1.35 (1.25 - 1.46) | 1.67 (1.43 - 1.95) |  |
| Angina pectoris | 386 (1.3) | 44 (2.1) | 722 (6.7) | 88 (8.2) | 1.74 (1.27 - 2.38) | 2.15 (1.88 - 2.47) | 2.86 (2.25 - 3.64) |  |
| **Pulmonary** |  |  |  |  |  |  |  |  |
| Respiratory failure | 606 (2.1) | 104 (5.2) | 1105 (10.4) | 171 (17.5) | 2.85 (2.31 - 3.52) | 1.62 (1.45 - 1.82) | 3.01 (2.51 - 3.60) | <sup>b</sup> |
| Pulmonary embolism | 259 (0.9) | 27 (1.3) | 363 (3.4) | 65 (6.1) | 1.57 (1.06 - 2.34) | 1.76 (1.47 - 2.11) | 3.33 (2.49 - 4.44) | <sup>a,b</sup> |
| Dyspnea | 5650 (20.2) | 814 (46.3) | 5688 (58.8) | 768 (100.7) | 2.32 (2.16 - 2.50) | 1.70 (1.63 - 1.77) | 3.06 (2.83 - 3.30) | <sup>a,b</sup> |
| **Neurological** |  |  |  |  |  |  |  |  |
| Nerve/nerve root/plexus disorder | 3005 (10.6) | 311 (15.4) | 2524 (24.2) | 337 (32.6) | 1.47 (1.30 - 1.65) | 1.55 (1.46 - 1.64) | 2.08 (1.85 - 2.34) | <sup>a,b</sup> |
| Anosmia (loss taste/smell) | 749 (2.6) | 123 (6.2) | 296 (2.7) | 69 (6.5) | 2.23 (1.85 - 2.70) | 1.14 (0.98 - 1.33) | 2.56 (1.98 - 3.30) | <sup>b</sup> |
| Other neurological | 156 (12.8) | 22 (10.4) | 198 (25.5) | 55 (36.9) | 0.88 (0.56 - 1.38) | 1.08 (0.84 - 1.39) | 1.66 (1.19 - 2.32) | <sup>b</sup> |
| Amnesia (loss of memories) | 1910 (6.7) | 207 (10.2) | 1949 (18.4) | 257 (24.7) | 1.59 (1.38 - 1.84) | 1.33 (1.24 - 1.44) | 1.99 (1.73 - 2.28) | <sup>b</sup> |
| **Menta behavioral** |  |  |  |  |  |  |  |  |
| Anxiety disorder | 16474 (65.7) | 1574 (93.5) | 6609 (76.3) | 840 (103.4) | 1.34 (1.27 - 1.41) | 1.20 (1.16 - 1.24) | 1.51 (1.41 - 1.62) | <sup>b</sup> |
| Mood disorder | 9168 (34.4) | 867 (46.7) | 4529 (48.7) | 547 (60.7) | 1.29 (1.20 - 1.38) | 1.43 (1.38 - 1.49) | 1.69 (1.55 - 1.85) | <sup>a,b</sup> |
| Psychotic disorder | 235 (0.8) | 20 (1.0) | 146 (1.3) | 24 (2.2) | 1.15 (0.73 - 1.82) | 1.28 (0.99 - 1.65) | 1.92 (1.22 - 3.03) |  |
| Substance misuse | 5567 (19.9) | 417 (21.2) | 2870 (28.7) | 230 (22.6) | 1.04 (0.94 - 1.15) | 1.27 (1.20 - 1.33) | 0.94 (0.82 - 1.08) | <sup>b</sup> |
| Sleep disorder, apnea | 7599 (27.7) | 809 (42.3) | 5679 (65.3) | 796 (97.4) | 1.54 (1.44 - 1.66) | 1.69 (1.62 - 1.75) | 2.46 (2.28 - 2.65) | <sup>a,b</sup> |
| **Musculoskeletal** |  |  |  |  |  |  |  |  |
| Joint pain | 19260 (74.4) | 1825 (102.7) | 12103 (143.3) | 1406 (175.2) | 1.40 (1.33 - 1.47) | 1.35 (1.32 - 1.39) | 1.68 (1.58 - 1.77) | <sup>a,b</sup> |
| Dorsalgia (back pain) | 18184 (71.6) | 1790 (104.8) | 10301 (122.9) | 1218 (155.7) | 1.46 (1.39 - 1.53) | 1.33 (1.30 - 1.37) | 1.67 (1.57 - 1.77) | <sup>a,b</sup> |
| Myalgia | 3258 (11.5) | 350 (18.1) | 1789 (16.9) | 238 (23.4) | 1.50 (1.35 - 1.68) | 1.20 (1.13 - 1.29) | 1.56 (1.36 - 1.79) | <sup>b</sup> |
| **General / Other** |  |  |  |  |  |  |  |  |
| Fatigue | 11067 (40.6) | 1338 (75.9) | 6196 (64.2) | 928 (106.9) | 1.79 (1.69 - 1.89) | 1.23 (1.19 - 1.28) | 1.95 (1.82 - 2.09) | <sup>b</sup> |
| Liver abnormality | 2987 (10.5) | 311 (15.4) | 3157 (30.7) | 369 (36.6) | 1.47 (1.31 - 1.66) | 2.02 (1.91 - 2.13) | 2.35 (2.10 - 2.62) | <sup>a,b</sup> |
| Kidney injury | 1592 (5.5) | 111 (5.5) | 2994 (30.2) | 250 (25.8) | 1.20 (0.99 - 1.45) | 2.35 (2.20 - 2.52) | 2.53 (2.20 - 2.90) | <sup>a</sup> |
| Atopic dermatitis | 1735 (6.1) | 157 (7.7) | 809 (7.5) | 52 (4.8) | 1.20 (1.02 - 1.41) | 1.11 (1.01 - 1.23) | 0.68 (0.51 - 0.90) | <sup>a,b</sup> |

<sup>a</sup>Significant difference compared with COVID-19 alone (p<0.05); p-values adjusted for multiple comparison with Holm's sequential Bonferroni procedure.
<sup>b</sup>Significant difference compared with any metabolic condition alone (p<0.05); p-values adjusted for multiple comparison with Holm's sequential Bonferroni procedure.
CI: confidence interval. Any metabolic condition included at least one of three baseline metabolic disorder (diabetes, hypertension, obesity) diagnosed between January 1, 2019 and index date. Mutually exclusive joint-exposure categories were based on baseline metabolic condition (yes/no) and COVID-19 diagnosis (yes/no), classifying participants as having neither (referent), COVID-19 only, baseline metabolic condition only, and both COVID-19 diagnosis and baseline metabolic condition. Models were adjusted for age (18-24, 25-34, 35-44, 45-54, 55-64, 65+), sex (male, female), region (northeast, north central, south, west), diagnosis time (January-March 2020, April-June 2020, July-September 2020, October-December 2020, January-March 2021, April-July 2021), and comorbidities (congestive heart failure, cardiac arrhythmias, valvular disease, pulmonary circulation disorders, peripheral vascular disorders, paralysis, other neurological disorders, chronic pulmonary disease, hypothyroidism, renal failure, liver disease, peptic ulcer disease, HIV/AIDS, lymphoma, metastatic cancer, solid tumor without metastasis, rheumatoid arthritis, coagulopathy, weight loss, fluid and electrolyte disorder, blood loss anemia, deficiency anemia, alcohol abuse, drug abuse, psychoses, depression, myocardial infraction, cerebrovascular disease, dementia, hemiplegia/paraplegia).

For almost all outcomes, adjusted hazard ratios were highest for the dual-exposed group (**Table 2** and **Figure 3**). Compared to COVID-19 patients without baseline metabolic conditions, the dual-exposed group had higher risks of developing almost all post-acute conditions and symptoms (HRs ranging from 1.51 to 3.33), except for substance misuse. The associations appeared strongest for pulmonary embolism (HR, 3.33; 95% CI, 2.49, 4.44), heart failure (HR, 3.09; 95% CI, 2.59, 3.68), dyspnea (HR, 3.06; 95% CI, 2.83, 3.30), and respiratory failure (HR, 3.01; 95% CI, 2.51, 3.60).

**Figure 3.**
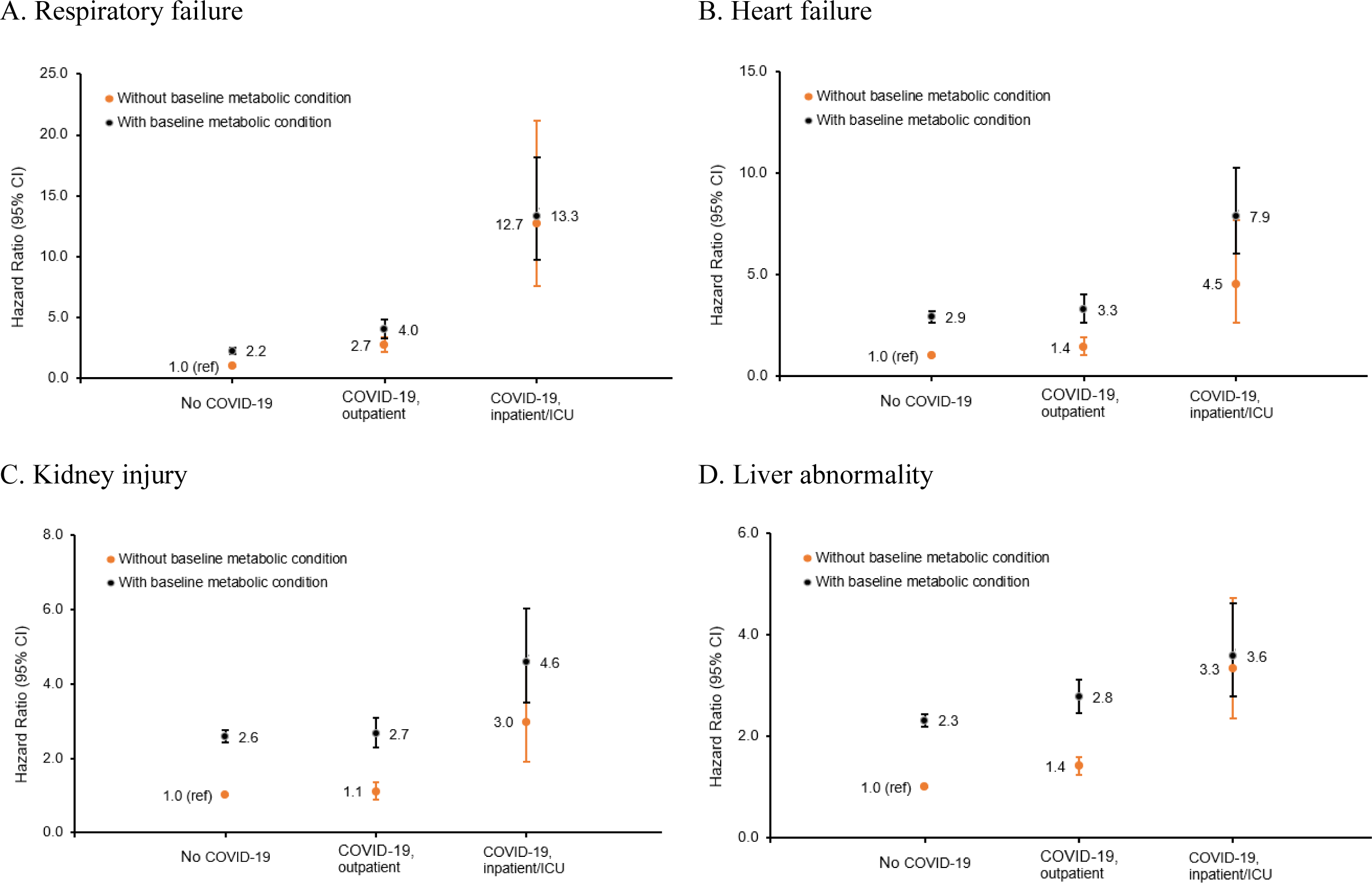
Multivariable adjusted hazard ratios for selected post-acute sequelae of COVID-19. Baseline metabolic conditions included obesity, diabetes, and/or hypertension, diagnosed between January 1, 2019 and index date. Models were adjusted for age (18-24, 25-34, 35-44, 45-54, 55-64, 65+), sex (male, female), region (northeast, north central, south, west), COVID-19 diagnosis time period (January-March 2020, April-June 2020, July-September 2020, October-December 2020, January-March 2021, April-July 2021), and comorbidities (congestive heart failure, cardiac arrhythmias, valvular disease, pulmonary circulation disorders, peripheral vascular disorders, paralysis, other neurological disorders, chronic pulmonary disease, hypothyroidism, renal failure, liver disease, peptic ulcer disease, HIV/AIDS, lymphoma, metastatic cancer, solid tumor without metastasis, rheumatoid arthritis, coagulopathy, weight loss, fluid and electrolyte disorder, blood loss anemia, deficiency anemia, alcohol abuse, drug abuse, psychoses, depression, myocardial infraction, cerebrovascular disease, dementia, hemiplegia/paraplegia).

Among adults without a baseline metabolic condition, the associations of COVID-19 with developing new symptoms and conditions were generally weaker, although the risks were still elevated for most conditions. Compared to individuals without a baseline metabolic condition or COVID-19 diagnosis, COVID-19 patients with a baseline metabolic condition had higher risks of developing respiratory failure (HR, 2.85; 95% CI, 2.31, 3.52), dyspnea (HR, 2.32; 95% CI, 2.16, 2.50), anosmia (HR, 2.23; 95% CI, 1.85, 2.70), and tachycardia (HR, 2.02; 95% CI, 1.80, 2.27). They also had 34% to 85% higher risk of developing other conditions and symptoms including chest pain, arrhythmia, fatigue, angina, heart failure, amnesia, sleep disorder, myalgia, liver abnormality, nerve disorders, dorsalgia, joint pain, and anxiety disorder compared to those without existing metabolic conditions.

The coefficients for the dual-exposed group were approximately the product of the HRs of having either condition alone, indicating an independent relationship between COVID-19 diagnosis and baseline metabolic condition. Post-hoc tests for the interaction term between COVID-19 diagnosis and baseline metabolic condition yielded mostly insignificant results. Post-hoc tests of the differences in coefficients indicated that among adults with a baseline metabolic condition, having COVID-19 was associated with higher risk of developing most of the symptoms and conditions and conversely that having an existing metabolic condition increased the risks of sequelae among COVID-19 patients.

**Table 3** presents the relationships between COVID-19 severity, baseline metabolic condition and risks of developing new symptoms and diseases, with outcomes illustrated in **Figure 3**. Among participants without a baseline metabolic condition, having a COVID-19 diagnosis was associated with increased risk of developing a new symptom or disease (HRs ranging from 0.89 to 2.72); those who had an inpatient and/or ICU admission had the highest risks (HRs ranging from 0.52 to 12.68). Similar patterns were observed among participants who had a baseline metabolic condition.

**Table 3.** Rates and hazard ratios for post-acute sequelae of COVID-19, stratified by COVID-19 severity.

| Outcomes | Without baseline metabolic condition, HR (95% CI) |  |  | With baseline metabolic condition, HR (95% CI) |  |  |
| --- | --- | --- | --- | --- | --- | --- |
|  | No COVID-19 | COVID-19, outpatient | COVID-19, inpatient/ICU | No COVID-19 | COVID-19, outpatient | COVID-19, inpatient/ICU |
| **Cardiovascular** |  |  |  |  |  |  |
| Heart failure | 1.00 [reference] | 1.41 (1.04 - 1.93) | 4.54 (2.67 - 7.71) | 2.93 (2.66 - 3.23) | 3.27 (2.66 - 4.02) | 7.88 (6.04 - 10.27) |
| Myocardial infraction | 1.00 [reference] | 1.04 (0.72 - 1.50) | 4.22 (2.32 - 7.67) | 2.18 (1.95 - 2.44) | 2.55 (1.99 - 3.25) | 3.69 (2.46 - 5.53) |
| Stroke | 1.00 [reference] | 1.10 (0.72 - 1.67) | 1.51 (0.49 - 4.71) | 2.09 (1.83 - 2.39) | 2.37 (1.77 - 3.19) | 5.61 (3.80 - 8.26) |
| Chest pain | 1.00 [reference] | 1.86 (1.74 - 2.00) | 2.74 (2.11 - 3.56) | 1.79 (1.72 - 1.85) | 2.63 (2.43 - 2.84) | 4.32 (3.66 - 5.10) |
| Deep vein thrombosis | 1.00 [reference] | 1.23 (0.91 - 1.68) | 5.51 (3.17 - 9.56) | 1.75 (1.55 - 1.98) | 2.49 (1.93 - 3.22) | 7.33 (5.25 - 10.24) |
| Tachycardia | 1.00 [reference] | 1.94 (1.72 - 2.19) | 5.78 (4.12 - 8.12) | 2.07 (1.94 - 2.22) | 2.79 (2.42 - 3.21) | 7.68 (6.10 - 9.66) |
| Arrhythmia | 1.00 [reference] | 1.81 (1.52 - 2.14) | 3.05 (1.98 - 4.70) | 1.57 (1.46 - 1.69) | 1.81 (1.52 - 2.16) | 3.29 (2.51 - 4.33) |
| Angina pectoris | 1.00 [reference] | 1.69 (1.22 - 2.35) | 2.96 (1.22 - 7.16) | 2.60 (2.27 - 2.97) | 3.44 (2.65 - 4.46) | 4.72 (2.96 - 7.51) |
| **Pulmonary** |  |  |  |  |  |  |
| Respiratory failure | 1.00 [reference] | 2.72 (2.17 - 3.40) | 12.68 (7.59 - 21.18) | 2.24 (2.02 - 2.50) | 4.01 (3.31 - 4.87) | 13.33 (9.77 - 18.19) |
| Pulmonary embolism | 1.00 [reference] | 1.30 (0.83 - 2.05) | 6.65 (3.13 - 14.12) | 2.18 (1.84 - 2.60) | 3.30 (2.35 - 4.64) | 10.78 (7.09 - 16.39) |
| Dyspnea | 1.00 [reference] | 2.35 (2.18 - 2.53) | 5.20 (3.81 - 7.10) | 2.03 (1.95 - 2.11) | 3.41 (3.14 - 3.70) | 9.49 (7.88 - 11.43) |
| **Neurological** |  |  |  |  |  |  |
| Nerve/nerve root/plexus disorder | 1.00 [reference] | 1.46 (1.29 - 1.64) | 2.30 (1.52 - 3.46) | 1.72 (1.63 - 1.83) | 2.47 (2.19 - 2.79) | 2.48 (1.86 - 3.32) |
| Anosmia (loss taste/smell) | 1.00 [reference] | 2.33 (1.93 - 2.83) | 0.52 (0.07 - 3.73) | 1.23 (1.06 - 1.42) | 2.92 (2.25 - 3.79) | 1.79 (0.74 - 4.32) |
| Other neurological | 1.00 [reference] | 1.49 (0.99 - 2.26) | 9.50 (5.83 - 15.47) | 1.56 (1.36 - 1.79) | 2.66 (2.01 - 3.53) | 8.59 (6.38 - 11.58) |
| Amnesia (loss of memories) | 1.00 [reference] | 1.62 (1.39 - 1.88) | 3.15 (2.07 - 4.80) | 1.62 (1.51 - 1.74) | 2.12 (1.82 - 2.47) | 5.23 (4.15 - 6.59) |
| **Menta behavioral** |  |  |  |  |  |  |
| Anxiety disorder | 1.00 [reference] | 1.36 (1.29 - 1.43) | 1.30 (0.99 - 1.71) | 1.37 (1.33 - 1.41) | 1.71 (1.58 - 1.84) | 2.32 (1.94 - 2.77) |
| Mood disorder | 1.00 [reference] | 1.30 (1.21 - 1.40) | 1.79 (1.32 - 2.42) | 1.59 (1.53 - 1.66) | 1.86 (1.70 - 2.05) | 2.50 (2.00 - 3.12) |
| Psychotic disorder | 1.00 [reference] | 0.89 (0.52 - 1.53) | 8.11 (3.59 - 18.34) | 1.67 (1.33 - 2.11) | 2.17 (1.30 - 3.62) | 7.19 (3.52 - 14.70) |
| Substance misuse | 1.00 [reference] | 1.05 (0.95 - 1.17) | 1.30 (0.84 - 1.99) | 1.39 (1.33 - 1.46) | 1.14 (0.99 - 1.31) | 0.85 (0.56 - 1.29) |
| Sleep disorder, apnea | 1.00 [reference] | 1.54 (1.43 - 1.66) | 2.45 (1.89 - 3.18) | 1.90 (1.83 - 1.98) | 2.77 (2.55 - 3.00) | 4.74 (4.03 - 5.56) |
| **Musculoskeletal** |  |  |  |  |  |  |
| Joint pain | 1.00 [reference] | 1.42 (1.35 - 1.49) | 1.45 (1.18 - 1.80) | 1.46 (1.42 - 1.49) | 1.90 (1.80 - 2.02) | 1.92 (1.66 - 2.22) |
| Dorsalgia (back pain) | 1.00 [reference] | 1.49 (1.42 - 1.57) | 1.27 (1.00 - 1.61) | 1.44 (1.41 - 1.48) | 1.90 (1.78 - 2.02) | 1.87 (1.59 - 2.20) |
| Myalgia | 1.00 [reference] | 1.54 (1.37 - 1.72) | 1.69 (1.02 - 2.81) | 1.41 (1.32 - 1.50) | 1.93 (1.68 - 2.22) | 2.01 (1.40 - 2.88) |
| **General / Other** |  |  |  |  |  |  |
| Fatigue | 1.00 [reference] | 1.82 (1.71 - 1.93) | 2.17 (1.70 - 2.78) | 1.40 (1.36 - 1.45) | 2.25 (2.09 - 2.42) | 3.03 (2.56 - 3.57) |
| Liver abnormality | 1.00 [reference] | 1.42 (1.25 - 1.60) | 3.33 (2.35 - 4.72) | 2.31 (2.19 - 2.44) | 2.77 (2.46 - 3.12) | 3.58 (2.78 - 4.62) |
| Kidney injury | 1.00 [reference] | 1.11 (0.90 - 1.37) | 2.98 (1.91 - 4.63) | 2.58 (2.42 - 2.75) | 2.66 (2.29 - 3.09) | 4.59 (3.49 - 6.02) |
| Atopic dermatitis | 1.00 [reference] | 1.27 (1.08 - 1.49) | 0.20 (0.03 - 1.45) | 1.24 (1.13 - 1.36) | 0.74 (0.55 - 1.00) | 1.01 (0.50 - 2.02) |

### Secondary analyses and sensitivity analyses

We repeated the analyses with joint exposure group by COVID-19 diagnosis and each of the metabolic disorders separately, and the results are presented in **Supplementary Tables 2-4**. The overall patterns were consistent with the main results. There was a slightly stronger association between baseline diabetes and risk of developing new clinical sequelae compared with hypertension and obesity. Stratifying by age and sex, the associations appeared to be stronger for younger adults compared with those for older adults (**Supplementary Tables 5and 6**). The differences by sex were small. Sensitivity checks with PSM yielded similar results (**Supplementary Tables 7and 8**), although the magnitude of associations was somewhat weaker with PSM-adjusted HRs for COVID-19 patients with existing metabolic conditions.

## Discussion

In a large national cohort of commercially insured adults, we found excess risk for clinical sequelae after the acute phase associated with COVID-19 infection, with or without a baseline metabolic condition. We found no significant interaction between COVID-19 and baseline metabolic condition. Among COVID-19 patients, having a baseline metabolic condition increased the risk of developing new clinical sequelae, indicating that metabolic disease is an independent risk factor for experiencing long-term post-COVID conditions.

The evidence linking metabolic disorders with severe COVID-19 and acute adverse outcomes is strong,^10^ but how underlying metabolic disease may influence long-term outcomes after SARS-CoV-2 infection remains unclear. Several studies have reported higher incidence of cardiovascular disease and neuropsychological conditions among COVID-19 patients compared with those not infected by the virus or those infected with influenza or respiratory tract diseases.^17,18,24^ A few studies have compared the risk of developing new symptoms or conditions after COVID-19 infection by baseline metabolic condition, finding a 30% higher risk of hospital admission among those with obesity^12^ but no increased risk among those with diabetes.^15^ The present study is among the first to assess the independent and joint associations of having COVID-19 and a baseline metabolic condition on the risks of developing new clinical sequelae.

Previous studies have reported high prevalence of comorbidities among patients infected with SARS-CoV-2.^25^ Our results suggest that among COVID-19 patients, those with baseline metabolic disorders may have even higher rates of other pre-existing comorbidities. The higher prevalence of comorbidities may partially explain the higher risks of developing new clinical sequelae post-acute phase among those individuals who had a baseline metabolic condition.

COVID-19 and metabolic disorders are both multisystem diseases that affect blood vessels, nerves, lung, kidney, liver, and muscle.^26^ Diabetes and hypertension are common pre-existing comorbidities among COVID-19 patients. Gene expression of angiotensin-converting enzyme, used by SARS-CoV-2 to enter human cells, was high in patients with diabetes, which may contribute to the higher risk of SARS-CoV-2 infection and developing PASC.^27^ Inflammation and immune memory are other hypothesized mechanisms.^28^ Due to high concentrations of chemokines, adipokines, and proinflammatory cytokines, obesity is characterized by constant chronic inflammation, which decreases activation of macrophages during the course of infection and impairs humoral and cellular immune memory.^27^ The interplay between metabolic diseases and COVID-19 could result in increased disease severity, breakthrough infections following vaccination, risk of complications, and development of new-onset diabetes.^11^ Given the overall high burden of metabolic disorders in the US and globally, COVID-19 could have important implications on the healthcare system and resources.

### Limitations

While the large sample enabled the assessment of detailed clinical outcomes, using commercial claims data for epidemiological research has a few well-known limitations. First, ICD-10-CM diagnoses can be unreliable.^29^ The estimated incidence for symptoms reflected only those reported and recorded, likely representing more severe or disturbing symptoms. Individuals with asymptomatic COVID-19 or those with less severe forms of metabolic disorders may have been misclassified. Second, information on socioeconomic status and lifestyle factors was not available, such as race, ethnicity, educational attainment, smoking, and exercise. Thus, the estimations are subject to omitted variable bias. Third, people included in the analyses represent only commercially insured adults, and results may not be generalizable to those with public health insurance or uninsured individuals. It is possible that individuals in the sample may have higher health literacy with better access to testing and care. Fourth, confidence intervals were not adjusted for multiple comparisons; however, estimate precision was high due to the large sample size. Estimates were also robust to additional adjustment, including Bonferroni procedures.

## Conclusion

In a large national cohort of commercially insured adults, COVID-19 patients with a baseline metabolic condition had higher risks of developing new clinical sequelae post-acute infection phase compared those who had a baseline metabolic condition but no COVID-19 diagnosis and those who had a COVID-19 diagnosis but no baseline metabolic disorder. COVID-19 could enhance the high burden of metabolic disorders in the US and globally, and future research should further investigate the interplay of these conditions with COVID-19.

## Data Availability

All data produced in the present study are available upon reasonable request to the authors.

## ACKNOWLEDGMENTS

Conflict of Interest Disclosures:

Dr. Stokes reported receiving grants from Johnson & Johnson, Inc. outside of the submitted work. No other disclosures were reported.

## Funding/Support

We gratefully acknowledge receiving financial support through grants from the Robert Wood Johnson Foundation (77521); the National Institute on Aging (R01-AG060115-04S1); the W.K. Kellogg Foundation (P-6007864-2022); the National Science Foundation (CCF-2200052); and Swiss Re, Inc. The funders had no role in the design and conduct of the study; collection, management, analysis, and interpretation of the data; preparation, review, or approval of the manuscript; or decision to submit the manuscript for publication.

## Author Contributions

Wubin Xie and Andrew Stokes had full access to all the data in this study and take responsibility for the integrity of the data and the accuracy of the data analysis.

---

**Figure 1.**
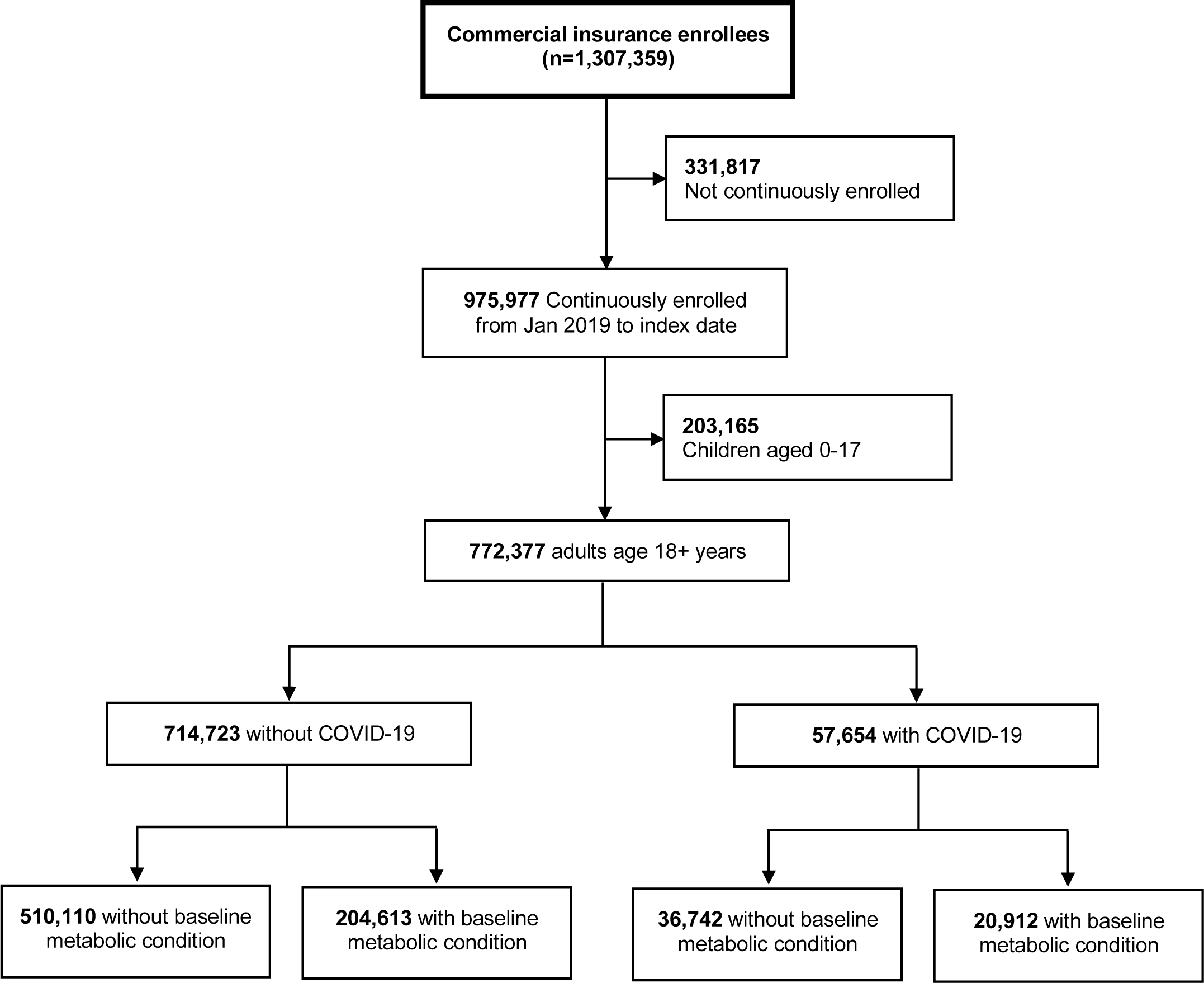
**Sample selection, IBM Watson MarketScan 2019-2021**

**Supplementary Table 1.** ICD-10-CM codes used to determine baseline metabolic conditions and incident outcomes.

| **Metabolic conditions** | **ICD-10-CM Codes** |
| --- | --- |
| 1. Diabetes | E10.0, E10.1, E10.9, E11.0, E11.1, E11.9, E12.0, E12.1, E12.9, E13.0, E13.1, E13.9, E14.0, E14.1, E14.9, E10.2 - E10.8, E11.2 - E11.8, E12.2 - E12.8, E13.2 - E13.8, E14.2 - E14.8 |
| 2. Hypertension | I10.x, I11.x - I13.x, I15.x |
| 3. Obesity | E66.x |
| **Outcome of interest** |  |
| *Cardiovascular* |  |
| 1. Heart failure | I50.x |
| 2. Myocardial infraction | I21.x-I22.x |
| 3. Stroke | I60.x-I64.x |
| 4. Chest pain | R07.9, R07.89 |
| 5. Deep vein thrombosis | I82 |
| 6. Tachycardia | I47.2, R00.0 |
| 7. Arrhythmia | I47-I49 |
| 8. Angina pectoris | I20.x |
| *Pulmonary* |  |
| 9. Respiratory failure | J96.0x, J961.x, J84* |
| 10. Pulmonary embolism | I26.0x, I26.9x |
| 11. Dyspnea | R06.0x |
| *Neurological* |  |
| 12. Nerve/nerve root/plexus disorder | G50.x-G59.x |
| 13. Loss taste or smell, anosmia | R43.x |
| 14. Other neurological conditions | G04.x, G05.x, A86.x, A85.8, G70.x-G73.x, G61.0, G20, G21, F01/F03, G30, G31.0, G31.83 |
| 15. Amnesia (loss of memories) | R41.x |
| *Mental behavioral* |  |
| 16. Anxiety disorder | F40.x-F48.x |
| 17. Mood disorder | F30.x-F39.x |
| 18. Psychotic disorder | F20.x-F29.x |
| 19. Substance misuse | F10.x-F19.x |
| 20. Sleep disorder, apnea | G47.x, F51.0 |
| *Musculoskeletal* |  |
| 21. Joint pain | M25.5x |
| 22. Dorsalgia (back pain) | M54.x |
| 23. Myalgia (muscle pain) | M79.1x |
| General / Other |  |
| 24. Fatigue | R53, R53.8x, G93.3 |
| 25. Kidney injury | N17.x, N18.x, I12.x, I13.x |
| 26. Liver abnormality | B18.x, K70, K70-K77 |
| 27. Atopic dermatitis (*negative control*) | L20x |

**Supplementary Table 2.** Rates and hazard ratios for post-acute sequelae of COVID-19, stratified by age group.

| Outcomes | Younger adults (Aged 18-64), Hazard Ratio (95% CI) |  |  |  |  |  | Older adults (Aged 18-64), Hazard Ratio (95% CI) |  |  |  |  |  |
| --- | --- | --- | --- | --- | --- | --- | --- | --- | --- | --- | --- | --- |
|  | COVID-19<br>vs None |  | Any metabolic<br>vs none |  | Both<br>vs none |  | COVID-19<br>vs None |  | Any metabolic<br>vs none |  | Both<br>vs none |  |
| Cardiovascular |  |  |  |  |  |  |  |  |  |  |  |  |
| Heart failure | 1.67 | (1.22 - 2.28) | 2.75 | (2.41 - 3.13) | 4.24 | (3.42 - 5.27) | 1.80 | (1.03 - 3.14) | 1.62 | (1.39 - 1.87) | 1.75 | (1.31 - 2.34) |
| Myocardial infraction | 1.31 | (0.93 - 1.85) | 2.08 | (1.80 - 2.40) | 2.86 | (2.20 - 3.71) | 1.15 | (0.51 - 2.61) | 1.25 | (1.04 - 1.52) | 0.93 | (0.62 - 1.39) |
| Stroke | 0.99 | (0.63 - 1.56) | 1.95 | (1.66 - 2.30) | 2.29 | (1.66 - 3.16) | 1.84 | (0.81 - 4.20) | 1.16 | (0.92 - 1.46) | 1.61 | (1.08 - 2.40) |
| Chest pain | 1.87 | (1.74 - 2.00) | 1.58 | (1.52 - 1.65) | 2.53 | (2.34 - 2.73) | 1.66 | (1.12 - 2.46) | 1.43 | (1.29 - 1.58) | 1.52 | (1.22 - 1.88) |
| Deep vein thrombosis | 1.38 | (1.03 - 1.85) | 1.50 | (1.30 - 1.73) | 2.47 | (1.92 - 3.18) | 2.29 | (1.06 - 4.95) | 1.06 | (0.83 - 1.37) | 1.78 | (1.16 - 2.73) |
| Tachycardia | 2.05 | (1.82 - 2.31) | 1.84 | (1.71 - 1.99) | 2.92 | (2.55 - 3.35) | 2.12 | (1.23 - 3.64) | 0.98 | (0.83 - 1.16) | 1.14 | (0.82 - 1.59) |
| Arrhythmia | 1.98 | (1.66 - 2.35) | 1.60 | (1.46 - 1.76) | 2.24 | (1.87 - 2.69) | 1.64 | (1.07 - 2.52) | 0.96 | (0.86 - 1.08) | 0.88 | (0.66 - 1.17) |
| Angina pectoris | 1.78 | (1.29 - 2.47) | 2.34 | (2.01 - 2.72) | 3.18 | (2.43 - 4.16) | 1.54 | (0.48 - 4.90) | 1.74 | (1.31 - 2.31) | 2.17 | (1.32 - 3.57) |
| Pulmonary |  |  |  |  |  |  |  |  |  |  |  |  |
| Respiratory failure | 2.95 | (2.34 - 3.71) | 1.98 | (1.73 - 2.27) | 3.78 | (3.04 - 4.69) | 3.17 | (1.89 - 5.29) | 1.04 | (0.87 - 1.24) | 1.70 | (1.26 - 2.30) |
| Pulmonary embolism | 1.28 | (0.81 - 2.03) | 1.92 | (1.57 - 2.36) | 3.86 | (2.78 - 5.36) | 4.83 | (2.17 - 10.73) | 1.43 | (1.01 - 2.02) | 2.28 | (1.29 - 4.04) |
| Dyspnea | 2.35 | (2.18 - 2.54) | 1.73 | (1.65 - 1.81) | 3.28 | (3.01 - 3.57) | 2.27 | (1.61 - 3.19) | 1.42 | (1.29 - 1.56) | 1.92 | (1.58 - 2.35) |
| Neurological |  |  |  |  |  |  |  |  |  |  |  |  |
| Nerve/nerve root/plexus disorder | 1.45 | (1.28 - 1.63) | 1.59 | (1.49 - 1.70) | 2.23 | (1.98 - 2.52) | 2.32 | (1.40 - 3.86) | 1.27 | (1.09 - 1.49) | 1.30 | (0.90 - 1.88) |
| Anosmia (loss taste/smell) | 2.28 | (1.88 - 2.76) | 1.20 | (1.03 - 1.40) | 2.41 | (1.83 - 3.16) | 0.00 | (0.00 - 0.00) | 0.75 | (0.45 - 1.23) | 3.92 | (1.88 - 8.18) |
| Other neurological | 0.94 | (0.59 - 1.49) | 1.11 | (0.84 - 1.47) | 1.92 | (1.33 - 2.77) | 0.56 | (0.07 - 4.16) | 0.94 | (0.58 - 1.52) | 1.08 | (0.52 - 2.23) |
| Amnesia (loss of memories) | 1.71 | (1.47 - 1.98) | 1.42 | (1.30 - 1.55) | 2.18 | (1.85 - 2.58) | 1.24 | (0.76 - 2.01) | 0.96 | (0.85 - 1.08) | 1.27 | (1.01 - 1.60) |
| Menta behavioral |  |  |  |  |  |  |  |  |  |  |  |  |
| Anxiety disorder | 1.35 | (1.28 - 1.42) | 1.22 | (1.18 - 1.26) | 1.52 | (1.41 - 1.64) | 1.05 | (0.67 - 1.64) | 1.01 | (0.92 - 1.12) | 1.32 | (1.06 - 1.64) |
| Mood disorder | 1.29 | (1.20 - 1.38) | 1.47 | (1.41 - 1.54) | 1.71 | (1.56 - 1.89) | 1.37 | (0.86 - 2.17) | 1.21 | (1.08 - 1.35) | 1.40 | (1.09 - 1.80) |
| Psychotic disorder | 0.78 | (0.45 - 1.37) | 1.27 | (0.96 - 1.69) | 1.66 | (0.92 - 3.00) | 9.39 | (3.85 - 22.89) | 1.43 | (0.82 - 2.49) | 2.58 | (1.17 - 5.68) |
| Substance misuse | 1.05 | (0.95 - 1.16) | 1.30 | (1.24 - 1.37) | 0.95 | (0.83 - 1.09) | 0.57 | (0.21 - 1.54) | 0.96 | (0.81 - 1.13) | 0.77 | (0.49 - 1.20) |
| Sleep disorder, apnea | 1.57 | (1.46 - 1.69) | 1.76 | (1.69 - 1.84) | 2.58 | (2.38 - 2.79) | 1.16 | (0.72 - 1.86) | 1.15 | (1.03 - 1.28) | 1.66 | (1.33 - 2.07) |
| Musculoskeletal |  |  |  |  |  |  |  |  |  |  |  |  |
| Joint pain | 1.40 | (1.34 - 1.47) | 1.37 | (1.34 - 1.41) | 1.72 | (1.62 - 1.83) | 1.45 | (1.10 - 1.91) | 1.17 | (1.10 - 1.26) | 1.37 | (1.17 - 1.61) |
| Dorsalgia (back pain) | 1.46 | (1.39 - 1.54) | 1.33 | (1.30 - 1.37) | 1.76 | (1.66 - 1.87) | 1.32 | (0.96 - 1.81) | 1.22 | (1.13 - 1.31) | 1.09 | (0.89 - 1.32) |
| Myalgia | 1.51 | (1.35 - 1.69) | 1.23 | (1.15 - 1.32) | 1.61 | (1.40 - 1.86) | 1.55 | (0.73 - 3.31) | 1.03 | (0.84 - 1.25) | 1.20 | (0.76 - 1.87) |
| General / Other |  |  |  |  |  |  |  |  |  |  |  |  |
| Fatigue | 1.79 | (1.69 - 1.90) | 1.21 | (1.16 - 1.25) | 1.99 | (1.85 - 2.14) | 1.72 | (1.15 - 2.58) | 1.30 | (1.17 - 1.44) | 1.74 | (1.41 - 2.14) |
| Liver abnormality | 1.48 | (1.31 - 1.66) | 2.17 | (2.05 - 2.30) | 2.52 | (2.24 - 2.83) | 2.17 | (1.27 - 3.73) | 1.15 | (0.98 - 1.35) | 1.35 | (0.96 - 1.92) |
| Kidney injury | 1.40 | (1.14 - 1.72) | 2.78 | (2.56 - 3.02) | 3.23 | (2.75 - 3.79) | 0.71 | (0.39 - 1.28) | 1.58 | (1.43 - 1.75) | 1.35 | (1.05 - 1.74) |
| Atopic dermatitis | 1.19 | (1.00 - 1.40) | 1.13 | (1.02 - 1.25) | 0.69 | (0.51 - 0.93) | 1.90 | (0.77 - 4.70) | 1.05 | (0.80 - 1.37) | 0.60 | (0.26 - 1.38) |

CI: confidence interval. COVID-19 group consisted of those diagnosed with COVID-19, but with no baseline metabolic condition. Any metabolic group consisted of those with a baseline metabolic condition, but no COVID-19 diagnosis. Baseline metabolic conditions included at least one of three baseline metabolic disorder (diabetes, hypertension, obesity) diagnosed between January 1, 2019 and index date. Models were adjusted for age (18-24, 25-34, 35-44, 45-54, 55-64, 65+), sex (male, female), region (northeast, north central, south, west), diagnosis time (January-March 2020, April-June 2020, July-September 2020, October-December 2020, January-March 2021, April-July 2021), and comorbidities (congestive heart failure, cardiac arrhythmias, valvular disease, pulmonary circulation disorders, peripheral vascular disorders, paralysis, other neurological disorders, chronic pulmonary disease, hypothyroidism, renal failure, liver disease, peptic ulcer disease, HIV/AIDS, lymphoma, metastatic cancer, solid tumor without metastasis, rheumatoid arthritis, coagulopathy, weight loss, fluid and electrolyte disorder, blood loss anemia, deficiency anemia, alcohol abuse, drug abuse, psychoses, depression, myocardial infraction, cerebrovascular disease, dementia, hemiplegia/paraplegia).

**Supplementary Table 3.** Rates and hazard ratios for post-acute sequelae of COVID-19, stratified by sex.

| Outcomes | Men, Hazard Ratio (95% CI) |  |  |  |  |  | Women, Hazard Ratio (95% CI) |  |  |  |  |  |
| --- | --- | --- | --- | --- | --- | --- | --- | --- | --- | --- | --- | --- |
|  | COVID-19<br>vs None |  | Any metabolic<br>vs none |  | Both<br>vs none |  | COVID-19<br>vs None |  | Any metabolic<br>vs none |  | Both<br>vs none |  |
| Cardiovascular |  |  |  |  |  |  |  |  |  |  |  |  |
| Heart failure | 2.15 | (1.54 - 3.01) | 2.08 | (1.81 - 2.38) | 3.09 | (2.43 - 3.93) | 1.06 | (0.67 - 1.69) | 2.41 | (2.10 - 2.77) | 3.07 | (2.39 - 3.93) |
| Myocardial infraction | 0.92 | (0.54 - 1.58) | 1.82 | (1.55 - 2.14) | 2.01 | (1.46 - 2.76) | 1.50 | (1.01 - 2.23) | 1.78 | (1.52 - 2.08) | 2.03 | (1.50 - 2.75) |
| Stroke | 1.27 | (0.79 - 2.02) | 1.51 | (1.28 - 1.80) | 2.03 | (1.47 - 2.80) | 0.77 | (0.36 - 1.64) | 1.96 | (1.59 - 2.42) | 2.48 | (1.68 - 3.66) |
| Chest pain | 1.73 | (1.15 - 1.92) | 1.63 | (1.55 - 1.72) | 2.46 | (2.21 - 2.75) | 1.95 | (1.79 - 2.12) | 1.53 | (1.46 - 1.61) | 2.35 | (2.13 - 2.59) |
| Deep vein thrombosis | 1.34 | (1.29 - 2.03) | 1.32 | (1.11 - 1.57) | 2.07 | (1.50 - 2.87) | 1.52 | (1.05 - 2.18) | 1.46 | (1.24 - 1.73) | 2.68 | (2.01 - 3.57) |
| Tachycardia | 1.86 | (1.51 - 2.28) | 2.12 | (1.92 - 2.35) | 3.17 | (2.62 - 3.84) | 2.09 | (1.81 - 2.40) | 1.45 | (1.32 - 1.58) | 2.21 | (1.88 - 2.61) |
| Arrhythmia | 1.65 | (0.84 - 2.09) | 1.36 | (1.23 - 1.50) | 1.94 | (1.58 - 2.38) | 2.03 | (1.64 - 2.53) | 1.31 | (1.18 - 1.46) | 1.41 | (1.12 - 1.78) |
| Angina pectoris | 1.78 | (0.88 - 2.73) | 2.14 | (1.78 - 2.57) | 2.75 | (1.95 - 3.87) | 1.69 | (1.08 - 2.66) | 2.36 | (1.95 - 2.86) | 3.45 | (2.48 - 4.78) |
| Pulmonary |  |  |  |  |  |  |  |  |  |  |  |  |
| Respiratory failure | 2.31 | (1.65 - 3.22) | 1.64 | (1.40 - 1.90) | 3.20 | (2.49 - 4.11) | 3.27 | (2.50 - 4.27) | 1.61 | (1.38 - 1.86) | 2.83 | (2.21 - 3.62) |
| Pulmonary embolism | 1.48 | (0.84 - 2.62) | 1.52 | (1.19 - 1.93) | 3.83 | (2.65 - 5.54) | 1.65 | (0.95 - 2.88) | 2.09 | (1.64 - 2.66) | 2.83 | (1.82 - 4.39) |
| Dyspnea | 2.08 | (1.84 - 2.35) | 1.80 | (1.69 - 1.91) | 3.10 | (2.76 - 3.49) | 2.49 | (2.27 - 2.73) | 1.59 | (1.51 - 1.68) | 2.93 | (2.65 - 3.24) |
| Neurological |  |  |  |  |  |  |  |  |  |  |  |  |
| Nerve/nerve root/plexus disorder | 1.31 | (1.06 - 1.61) | 1.78 | (1.63 - 1.95) | 2.68 | (2.25 - 3.19) | 1.55 | (1.35 - 1.79) | 1.45 | (1.34 - 1.56) | 1.94 | (1.67 - 2.26) |
| Anosmia (loss taste/smell) | 1.88 | (1.34 - 2.65) | 1.15 | (0.91 - 1.45) | 2.59 | (1.71 - 3.92) | 2.43 | (1.93 - 3.07) | 1.15 | (0.95 - 1.38) | 2.55 | (1.86 - 3.51) |
| Other neurological | 0.70 | (0.35 - 1.41) | 0.96 | (0.68 - 1.35) | 1.71 | (1.09 - 2.70) | 1.12 | (0.63 - 2.01) | 1.25 | (0.91 - 1.72) | 1.78 | (1.12 - 2.81) |
| Amnesia (loss of memories) | 1.64 | (1.31 - 2.07) | 1.34 | (1.21 - 1.49) | 1.79 | (1.44 - 2.21) | 1.58 | (1.32 - 1.90) | 1.18 | (1.08 - 1.30) | 1.69 | (1.43 - 2.01) |
| Menta behavioral |  |  |  |  |  |  |  |  |  |  |  |  |
| Anxiety disorder | 1.38 | (1.26 - 1.51) | 1.35 | (1.28 - 1.42) | 1.88 | (1.67 - 2.10) | 1.32 | (1.24 - 1.40) | 1.13 | (1.09 - 1.17) | 1.34 | (1.23 - 1.47) |
| Mood disorder | 1.44 | (1.27 - 1.63) | 1.56 | (1.46 - 1.67) | 1.94 | (1.67 - 2.25) | 1.22 | (1.12 - 1.33) | 1.38 | (1.32 - 1.45) | 1.54 | (1.38 - 1.72) |
| Psychotic disorder | 1.11 | (0.57 - 2.19) | 1.12 | (0.80 - 1.56) | 1.60 | (0.82 - 3.12) | 1.19 | (0.64 - 2.21) | 1.30 | (0.95 - 1.78) | 1.77 | (0.99 - 3.17) |
| Substance misuse | 1.13 | (0.99 - 1.29) | 1.33 | (1.25 - 1.42) | 1.14 | (0.96 - 1.35) | 0.94 | (0.81 - 1.09) | 1.23 | (1.14 - 1.32) | 0.81 | (0.66 - 1.00) |
| Sleep disorder, apnea | 1.43 | (1.28 - 1.60) | 1.83 | (1.73 - 1.92) | 2.84 | (2.55 - 3.16) | 1.64 | (1.49 - 1.81) | 1.60 | (1.52 - 1.68) | 2.40 | (2.16 - 2.66) |
| Musculoskeletal |  |  |  |  |  |  |  |  |  |  |  |  |
| Joint pain | 1.43 | (1.33 - 1.54) | 1.39 | (1.34 - 1.44) | 1.74 | (1.60 - 1.90) | 1.38 | (1.30 - 1.47) | 1.33 | (1.29 - 1.37) | 1.72 | (1.60 - 1.85) |
| Dorsalgia (back pain) | 1.39 | (1.29 - 1.51) | 1.45 | (1.40 - 1.51) | 1.83 | (1.67 - 2.00) | 1.49 | (1.41 - 1.59) | 1.24 | (1.20 - 1.29) | 1.62 | (1.50 - 1.75) |
| Myalgia | 1.48 | (1.22 - 1.80) | 1.43 | (1.30 - 1.58) | 1.55 | (1.23 - 1.97) | 1.51 | (1.32 - 1.73) | 1.10 | (1.01 - 1.19) | 1.63 | (1.38 - 1.92) |
| General / Other |  |  |  |  |  |  |  |  |  |  |  |  |
| Fatigue | 1.73 | (1.57 - 1.92) | 1.45 | (1.38 - 1.53) | 2.38 | (2.14 - 2.66) | 1.81 | (1.69 - 1.94) | 1.10 | (1.05 - 1.15) | 1.76 | (1.61 - 1.92) |
| Liver abnormality | 1.78 | (1.51 - 2.10) | 2.16 | (1.99 - 2.33) | 2.74 | (2.34 - 3.22) | 1.24 | (1.05 - 1.47) | 2.05 | (1.91 - 2.21) | 2.45 | (2.11 - 2.86) |
| Kidney injury | 1.38 | (1.07 - 1.78) | 2.23 | (2.04 - 2.43) | 2.48 | (2.05 - 2.99) | 1.02 | (0.76 - 1.37) | 2.32 | (2.11 - 2.55) | 2.43 | (2.00 - 2.95) |
| Atopic dermatitis | 1.05 | (0.76 - 1.43) | 1.44 | (1.25 - 1.66) | 0.80 | (0.50 - 1.26) | 1.26 | (1.04 - 1.52) | 0.97 | (0.86 - 1.09) | 0.62 | (0.43 - 0.87) |

CI: confidence interval. COVID-19 group consisted of those diagnosed with COVID-19, but with no baseline metabolic condition. Any metabolic group consisted of those with a baseline metabolic condition, but no COVID-19 diagnosis. Baseline metabolic conditions included at least one of three baseline metabolic disorder (diabetes, hypertension, obesity) diagnosed between January 1, 2019 and index date. Models were adjusted for age (18-24, 25-34, 35-44, 45-54, 55-64, 65+), sex (male, female), region (northeast, north central, south, west), diagnosis time (January-March 2020, April-June 2020, July-September 2020, October-December 2020, January-March 2021, April-July 2021), and comorbidities (congestive heart failure, cardiac arrhythmias, valvular disease, pulmonary circulation disorders, peripheral vascular disorders, paralysis, other neurological disorders, chronic pulmonary disease, hypothyroidism, renal failure, liver disease, peptic ulcer disease, HIV/AIDS, lymphoma, metastatic cancer, solid tumor without metastasis, rheumatoid arthritis, coagulopathy, weight loss, fluid and electrolyte disorder, blood loss anemia, deficiency anemia, alcohol abuse, drug abuse, psychoses, depression, myocardial infraction, cerebrovascular disease, dementia, hemiplegia/paraplegia).

**Supplementary Table 4.** Rates and hazard ratios for post-acute sequelae of COVID-19 by baseline diabetes and COVID-19 infection.

| Outcomes | # of cases (Incidence per 1,000 person-years) |  |  |  | Adjusted Hazard Ratio (95% CI) |  |  |  |  |
| --- | --- | --- | --- | --- | --- | --- | --- | --- | --- |
|  | None | COVID-19 | Diabetes | Both | COVID-19 vs None |  | Diabetes vs none |  | Both vs none |
| **Cardiovascular** |  |  |  |  |  |  |  |  |  |
| Heart failure | 1580 (4.4) | 152 (5.5) | 690 (22.4) | 75 (23.2) | 1.42 (1.20 - 1.68) |  | 1.55 (1.41 - 1.71) |  | 2.18 (1.72 - 2.75) |
| Myocardial infraction | 1118 (3.1) | 88 (3.2) | 411 (12.8) | 52 (15.3) | 1.10 (0.88 - 1.36) |  | 1.51 (1.34 - 1.71) |  | 1.84 (1.38 - 2.45) |
| Stroke | 791 (2.2) | 64 (2.3) | 321 (10.0) | 41 (12.0) | 1.13 (0.88 - 1.47) |  | 1.56 (1.35 - 1.80) |  | 2.03 (1.46 - 2.80) |
| Chest pain | 11754 (33.9) | 1519 (61.6) | 2026 (70.2) | 285 (103.6) | 1.73 (1.64 - 1.82) |  | 1.24 (1.18 - 1.30) |  | 1.76 (1.56 - 1.99) |
| Deep vein thrombosis | 997 (2.7) | 123 (4.4) | 239 (7.4) | 37 (10.8) | 1.60 (1.32 - 1.93) |  | 1.17 (1.00 - 1.36) |  | 1.71 (1.22 - 2.40) |
| Tachycardia | 3487 (9.7) | 533 (19.7) | 678 (21.5) | 97 (30.2) | 1.89 (1.72 - 2.07) |  | 1.38 (1.26 - 1.51) |  | 1.80 (1.46 - 2.21) |
| Arrhythmia | 2712 (7.6) | 279 (10.2) | 681 (22.5) | 78 (24.1) | 1.53 (1.35 - 1.74) |  | 1.15 (1.05 - 1.26) |  | 1.44 (1.14 - 1.81) |
| Angina pectoris | 820 (2.3) | 98 (3.5) | 288 (8.9) | 34 (9.9) | 1.56 (1.26 - 1.93) |  | 1.43 (1.24 - 1.66) |  | 1.64 (1.15 - 2.32) |
| **Pulmonary** |  |  |  |  |  |  |  |  |  |
| Respiratory failure | 1243 (3.4) | 199 (7.4) | 468 (14.7) | 76 (25.5) | 2.27 (1.95 - 2.65) |  | 1.49 (1.33 - 1.67) |  | 2.75 (2.16 - 3.49) |
| Pulmonary embolism | 498 (1.4) | 70 (2.5) | 124 (3.8) | 22 (6.4) | 1.80 (1.39 - 2.32) |  | 1.10 (0.89 - 1.36) |  | 1.90 (1.22 - 2.95) |
| Dyspnea | 9386 (26.9) | 1321 (57.7) | 1952 (68.1) | 261 (113.3) | 2.10 (1.98 - 2.23) |  | 1.26 (1.20 - 1.33) |  | 2.22 (1.96 - 2.52) |
| **Neurological** |  |  |  |  |  |  |  |  |  |
| Nerve/nerve root/plexus disorder | 4685 (13.1) | 533 (19.6) | 844 (27.0) | 115 (34.6) | 1.43 (1.31 - 1.57) |  | 1.26 (1.16 - 1.36) |  | 1.56 (1.29 - 1.89) |
| Anosmia (loss taste/smell) | 961 (2.6) | 180 (6.7) | 84 (2.6) | 12 (3.5) | 2.34 (2.00 - 2.75) |  | 1.05 (0.83 - 1.34) |  | 1.35 (0.76 - 2.41) |
| Other neurological | 267 (15.5) | 53 (17.6) | 87 (33.0) | 24 (41.0) | 1.24 (0.92 - 1.67) |  | 1.23 (0.94 - 1.61) |  | 1.60 (1.03 - 2.49) |
| Amnesia (loss of memories) | 3137 (8.7) | 372 (13.6) | 722 (22.8) | 92 (28.0) | 1.59 (1.42 - 1.77) |  | 1.28 (1.17 - 1.40) |  | 1.76 (1.42 - 2.17) |
| **Menta behavioral** |  |  |  |  |  |  |  |  |  |
| Anxiety disorder | 21213 (68.4) | 2154 (97.0) | 1870 (68.2) | 260 (94.5) | 1.32 (1.26 - 1.38) |  | 0.97 (0.92 - 1.02) |  | 1.25 (1.11 - 1.42) |
| Mood disorder | 12318 (37.2) | 1220 (49.5) | 1379 (48.9) | 194 (66.6) | 1.24 (1.16 - 1.31) |  | 1.18 (1.11 - 1.25) |  | 1.53 (1.33 - 1.77) |
| Psychotic disorder | 338 (0.9) | 31 (1.1) | 43 (1.3) | 13 (3.7) | 1.09 (0.75 - 1.58) |  | 0.96 (0.67 - 1.36) |  | 2.57 (1.43 - 4.60) |
| Substance misuse | 7576 (21.7) | 573 (21.5) | 861 (28.4) | 74 (22.7) | 0.95 (0.87 - 1.03) |  | 1.06 (0.98 - 1.14) |  | 0.79 (0.63 - 1.00) |
| Sleep disorder, apnea | 11505 (34.3) | 1331 (53.9) | 1773 (69.4) | 274 (106.0) | 1.51 (1.42 - 1.59) |  | 1.19 (1.12 - 1.25) |  | 1.71 (1.51 - 1.93) |
| **Musculoskeletal** |  |  |  |  |  |  |  |  |  |
| Joint pain | 27794 (87.4) | 2792 (120.4) | 3569 (139.5) | 439 (169.1) | 1.35 (1.30 - 1.40) |  | 1.02 (0.98 - 1.06) |  | 1.23 (1.12 - 1.35) |
| Dorsalgia (back pain) | 25368 (81.2) | 2605 (116.6) | 3117 (122.7) | 403 (157.8) | 1.39 (1.33 - 1.45) |  | 1.07 (1.03 - 1.12) |  | 1.34 (1.22 - 1.49) |
| Myalgia | 4539 (12.7) | 515 (19.6) | 508 (16.0) | 73 (22.2) | 1.44 (1.31 - 1.57) |  | 1.01 (0.91 - 1.11) |  | 1.30 (1.03 - 1.64) |
| **General / Other** |  |  |  |  |  |  |  |  |  |
| Fatigue | 15537 (45.8) | 1991 (84.9) | 1726 (58.8) | 275 (96.2) | 1.73 (1.65 - 1.81) |  | 0.95 (0.90 - 1.00) |  | 1.46 (1.29 - 1.65) |
| Liver abnormality | 5010 (14.0) | 547 (20.2) | 1134 (37.2) | 133 (42.1) | 1.36 (1.24 - 1.48) |  | 1.51 (1.40 - 1.62) |  | 1.61 (1.35 - 1.91) |
| Kidney injury | 3311 (9.3) | 245 (9.0) | 1275 (45.0) | 116 (39.9) | 1.11 (0.97 - 1.27) |  | 1.82 (1.70 - 1.95) |  | 1.98 (1.64 - 2.39) |
| Atopic dermatitis | 2313 (6.4) | 200 (7.2) | 231 (7.1) | 9 (2.6) | 1.06 (0.91 - 1.22) |  | 1.00 (0.86 - 1.15) |  | 0.35 (0.18 - 0.67) |

CI: confidence interval. Existing hypertension was between January 1, 2019 and index date. Models were adjusted for age (18-24, 25-34, 35-44, 45-54, 55-64, 65+), sex (male, female), region (northeast, north central, south, west), diagnosis time (January-March 2020, April-June 2020, July-September 2020, October-December 2020, January-March 2021, April-July 2021), and comorbidities (congestive heart failure, cardiac arrhythmias, valvular disease, pulmonary circulation disorders, peripheral vascular disorders, paralysis, other neurological disorders, chronic pulmonary disease, hypothyroidism, renal failure, liver disease, peptic ulcer disease, HIV/AIDS, lymphoma, metastatic cancer, solid tumor without metastasis, rheumatoid arthritis, coagulopathy, weight loss, fluid and electrolyte disorder, blood loss anemia, deficiency anemia, alcohol abuse, drug abuse, psychoses, depression, myocardial infraction, cerebrovascular disease, dementia, hemiplegia/paraplegia).

**Supplementary Table 5.** Rates and hazard ratios for post-acute sequelae of COVID-19 by baseline hypertension and COVID-19 infection.

| Outcomes | # of cases (Incidence per 1,000 person-years) |  |  |  | Adjusted Hazard Ratio (95% CI) |  |  |
| --- | --- | --- | --- | --- | --- | --- | --- |
|  | None | COVID-19 | Hypertension | Both | COVID-19 vs None | Hypertension vs none | Both vs none |
| **Cardiovascular** |  |  |  |  |  |  |  |
| Heart failure | 787 (2.5) | 74 (3.2) | 1483 (18.7) | 153 (19.9) | 1.51 (1.19 - 1.92) | 1.94 (1.75 - 2.14) | 2.63 (2.19 - 3.15) |
| Myocardial infraction | 669 (2.1) | 58 (2.5) | 860 (10.5) | 82 (10.4) | 1.31 (1.00 - 1.71) | 1.48 (1.31 - 1.67) | 1.53 (1.20 - 1.95) |
| Stroke | 483 (1.5) | 35 (1.5) | 629 (7.6) | 70 (8.8) | 1.11 (0.79 - 1.57) | 1.40 (1.22 - 1.62) | 1.74 (1.33 - 2.27) |
| Chest pain | 8888 (29.4) | 1146 (54.7) | 4892 (66.9) | 658 (101.6) | 1.81 (1.70 - 1.93) | 1.46 (1.40 - 1.52) | 2.14 (1.97 - 2.32) |
| Deep vein thrombosis | 701 (2.2) | 75 (3.2) | 535 (6.5) | 85 (10.7) | 1.49 (1.18 - 1.89) | 1.16 (1.01 - 1.32) | 1.89 (1.49 - 2.41) |
| Tachycardia | 2618 (8.4) | 403 (17.7) | 1547 (19.2) | 227 (29.8) | 2.00 (1.80 - 2.22) | 1.52 (1.40 - 1.64) | 2.21 (1.91 - 2.55) |
| Arrhythmia | 1775 (5.7) | 186 (8.1) | 1618 (21.1) | 171 (22.9) | 1.68 (1.45 - 1.96) | 1.29 (1.19 - 1.40) | 1.65 (1.39 - 1.94) |
| Angina pectoris | 463 (1.5) | 55 (2.4) | 645 (7.8) | 77 (9.6) | 1.69 (1.28 - 2.24) | 1.93 (1.68 - 2.22) | 2.48 (1.92 - 3.20) |
| **Pulmonary** |  |  |  |  |  |  |  |
| Respiratory failure | 742 (2.4) | 126 (5.6) | 969 (11.9) | 149 (20.6) | 2.65 (2.19 - 3.20) | 1.35 (1.21 - 1.52) | 2.46 (2.04 - 2.98) |
| Pulmonary embolism | 321 (1.0) | 39 (1.7) | 301 (3.6) | 53 (6.6) | 1.70 (1.22 - 2.38) | 1.37 (1.14 - 1.66) | 2.53 (1.85 - 3.47) |
| Dyspnea | 6632 (21.8) | 968 (49.4) | 4706 (64.1) | 614 (109.7) | 2.25 (2.11 - 2.41) | 1.52 (1.45 - 1.59) | 2.68 (2.45 - 2.92) |
| **Neurological** |  |  |  |  |  |  |  |
| Nerve/nerve root/plexus disorder | 3551 (11.5) | 393 (17.3) | 1978 (24.7) | 255 (33.0) | 1.47 (1.32 - 1.63) | 1.30 (1.22 - 1.38) | 1.68 (1.47 - 1.92) |
| Anosmia (loss taste/smell) | 842 (2.7) | 140 (6.3) | 203 (2.4) | 52 (6.6) | 2.15 (1.80 - 2.58) | 0.98 (0.82 - 1.17) | 2.46 (1.83 - 3.30) |
| Other neurological | 192 (13.8) | 34 (13.9) | 162 (26.9) | 43 (37.2) | 1.08 (0.75 - 1.56) | 0.98 (0.76 - 1.28) | 1.43 (0.99 - 2.07) |
| Amnesia (loss of memories) | 2229 (7.2) | 252 (11.0) | 1630 (20.1) | 212 (27.4) | 1.57 (1.38 - 1.79) | 1.17 (1.08 - 1.27) | 1.76 (1.51 - 2.04) |
| **Menta behavioral** |  |  |  |  |  |  |  |
| Anxiety disorder | 18140 (67.1) | 1775 (94.1) | 4943 (73.7) | 639 (104.7) | 1.31 (1.25 - 1.38) | 1.18 (1.14 - 1.23) | 1.56 (1.44 - 1.70) |
| Mood disorder | 10351 (36.0) | 1000 (48.1) | 3346 (46.4) | 414 (61.2) | 1.25 (1.17 - 1.34) | 1.22 (1.17 - 1.28) | 1.49 (1.35 - 1.65) |
| Psychotic disorder | 263 (0.8) | 23 (1.0) | 118 (1.4) | 21 (2.6) | 1.14 (0.74 - 1.74) | 1.21 (0.91 - 1.61) | 1.96 (1.19 - 3.21) |
| Substance misuse | 6164 (20.4) | 464 (20.8) | 2273 (29.7) | 183 (24.2) | 1.00 (0.91 - 1.10) | 1.30 (1.23 - 1.38) | 0.99 (0.85 - 1.16) |
| Sleep disorder, apnea | 8843 (30.0) | 976 (45.8) | 4435 (66.8) | 629 (105.0) | 1.51 (1.41 - 1.61) | 1.39 (1.34 - 1.45) | 2.06 (1.89 - 2.24) |
| **Musculoskeletal** |  |  |  |  |  |  |  |
| Joint pain | 21878 (78.4) | 2147 (108.0) | 9485 (147.4) | 1084 (183.3) | 1.37 (1.31 - 1.43) | 1.23 (1.20 - 1.27) | 1.54 (1.44 - 1.64) |
| Dorsalgia (back pain) | 20501 (74.9) | 2067 (108.3) | 7984 (124.5) | 941 (162.1) | 1.42 (1.35 - 1.48) | 1.22 (1.19 - 1.26) | 1.56 (1.46 - 1.67) |
| Myalgia | 3728 (12.1) | 410 (18.7) | 1319 (16.2) | 178 (23.3) | 1.45 (1.31 - 1.61) | 1.07 (0.99 - 1.15) | 1.43 (1.22 - 1.67) |
| **General / Other** |  |  |  |  |  |  |  |
| Fatigue | 12487 (42.4) | 1551 (78.3) | 4776 (64.2) | 715 (110.1) | 1.75 (1.66 - 1.84) | 1.17 (1.13 - 1.22) | 1.89 (1.75 - 2.05) |
| Liver abnormality | 3701 (12.0) | 391 (17.2) | 2443 (30.9) | 289 (38.4) | 1.39 (1.25 - 1.55) | 1.47 (1.38 - 1.56) | 1.72 (1.52 - 1.95) |
| Kidney injury | 1994 (6.4) | 151 (6.6) | 2592 (34.7) | 210 (29.8) | 1.21 (1.03 - 1.43) | 1.86 (1.74 - 1.99) | 1.93 (1.67 - 2.24) |
| Atopic dermatitis | 1906 (6.1) | 171 (7.4) | 638 (7.7) | 38 (4.7) | 1.14 (0.97 - 1.33) | 1.19 (1.07 - 1.33) | 0.69 (0.50 - 0.96) |

CI: Confidence interval. Baseline hypertension was assessed as diagnosed between January 1, 2019 and index date. Models were adjusted for age (18-24, 25-34, 35-44, 45-54, 55-64, 65+), sex (male, female), region (northeast, north central, south, west), diagnosis time (January-March 2020, April-June 2020, July-September 2020, October-December 2020, January-March 2021, April-July 2021), and comorbidities (congestive heart failure, cardiac arrhythmias, valvular disease, pulmonary circulation disorders, peripheral vascular disorders, paralysis, other neurological disorders, chronic pulmonary disease, hypothyroidism, renal failure, liver disease, peptic ulcer disease, HIV/AIDS, lymphoma, metastatic cancer, solid tumor without metastasis, rheumatoid arthritis, coagulopathy, weight loss, fluid and electrolyte disorder, blood loss anemia, deficiency anemia, alcohol abuse, drug abuse, psychoses, depression, myocardial infraction, cerebrovascular disease, dementia, hemiplegia/paraplegia).

**Supplementary Table 6.** Rates and hazard ratios for post-acute sequelae of COVID-19 by baseline obesity and COVID-19 infection.

| Outcomes | # of cases (Incidence per 1,000 person-years) |  |  |  | Adjusted Hazard Ratio (95% CI) |  |  |
| --- | --- | --- | --- | --- | --- | --- | --- |
|  | None | COVID-19 | Obesity | Both | COVID-19<br>vs None | Obesity<br>vs none | Both<br>vs none |
| **Cardiovascular** |  |  |  |  |  |  |  |
| Heart failure | 1851 (5.2) | 171 (6.4) | 419 (12.2) | 56 (13.8) | 1.39 (1.19 - 1.63) | 1.12 (1.00 - 1.26) | 1.67 (1.27 - 2.19) |
| Myocardial infraction | 1289 (3.6) | 103 (3.8) | 240 (6.8) | 37 (8.9) | 1.08 (0.88 - 1.32) | 1.02 (0.88 - 1.18) | 1.36 (0.97 - 1.90) |
| Stroke | 916 (2.5) | 82 (3.0) | 196 (5.5) | 23 (5.6) | 1.24 (0.98 - 1.55) | 1.09 (0.92 - 1.28) | 1.15 (0.76 - 1.76) |
| Chest pain | 11799 (34.3) | 1482 (61.5) | 1981 (63.4) | 322 (96.7) | 1.72 (1.63 - 1.81) | 1.21 (1.15 - 1.27) | 1.80 (1.61 - 2.02) |
| Deep vein thrombosis | 1002 (2.8) | 117 (4.3) | 234 (6.6) | 43 (10.5) | 1.54 (1.27 - 1.87) | 1.46 (1.25 - 1.70) | 2.42 (1.77 - 3.30) |
| Tachycardia | 3532 (9.9) | 507 (19.3) | 633 (18.4) | 123 (31.1) | 1.81 (1.65 - 1.99) | 1.18 (1.08 - 1.29) | 1.88 (1.56 - 2.26) |
| Arrhythmia | 2909 (8.2) | 294 (11.1) | 484 (14.4) | 63 (16.0) | 1.51 (1.34 - 1.71) | 1.11 (1.00 - 1.22) | 1.40 (1.09 - 1.80) |
| Angina pectoris | 900 (2.5) | 97 (3.6) | 208 (5.9) | 35 (8.5) | 1.41 (1.14 - 1.74) | 1.19 (1.01 - 1.39) | 1.75 (1.24 - 2.46) |
| **Pulmonary** |  |  |  |  |  |  |  |
| Respiratory failure | 1382 (3.8) | 207 (8.0) | 329 (9.4) | 68 (17.9) | 2.19 (1.89 - 2.54) | 1.22 (1.08 - 1.39) | 2.44 (1.90 - 3.13) |
| Pulmonary embolism | 485 (1.3) | 65 (2.4) | 137 (3.9) | 27 (6.5) | 1.80 (1.39 - 2.34) | 1.74 (1.42 - 2.12) | 3.02 (2.02 - 4.53) |
| Dyspnea | 9485 (27.4) | 1305 (58.4) | 1853 (59.6) | 277 (96.7) | 2.13 (2.01 - 2.26) | 1.32 (1.25 - 1.39) | 2.22 (1.97 - 2.51) |
| **Neurological** |  |  |  |  |  |  |  |
| Nerve/nerve root/plexus disorder | 4607 (13.0) | 497 (18.7) | 922 (27.0) | 151 (38.1) | 1.40 (1.27 - 1.54) | 1.37 (1.27 - 1.48) | 1.90 (1.61 - 2.24) |
| Anosmia (loss taste/smell) | 915 (2.5) | 159 (6.1) | 130 (3.7) | 33 (8.1) | 2.26 (1.91 - 2.67) | 1.30 (1.07 - 1.58) | 2.74 (1.92 - 3.91) |
| Other neurological | 295 (17.3) | 52 (17.4) | 59 (20.6) | 25 (41.2) | 1.08 (0.80 - 1.46) | 0.88 (0.66 - 1.18) | 1.73 (1.13 - 2.65) |
| Amnesia (loss of memories) | 3296 (9.2) | 369 (13.8) | 563 (16.1) | 95 (23.4) | 1.53 (1.38 - 1.71) | 1.10 (1.00 - 1.21) | 1.73 (1.41 - 2.13) |
| **Menta behavioral** |  |  |  |  |  |  |  |
| Anxiety disorder | 20700 (66.7) | 2054 (93.4) | 2383 (88.3) | 360 (121.0) | 1.32 (1.26 - 1.38) | 1.10 (1.05 - 1.15) | 1.42 (1.27 - 1.57) |
| Mood disorder | 12033 (36.4) | 1175 (48.5) | 1664 (57.0) | 239 (71.4) | 1.26 (1.18 - 1.33) | 1.30 (1.23 - 1.38) | 1.54 (1.35 - 1.76) |
| Psychotic disorder | 334 (0.9) | 36 (1.3) | 47 (1.3) | 8 (1.9) | 1.29 (0.91 - 1.83) | 1.15 (0.83 - 1.59) | 1.66 (0.81 - 3.39) |
| Substance misuse | 7598 (21.9) | 563 (21.7) | 839 (25.6) | 84 (21.3) | 0.94 (0.87 - 1.03) | 0.94 (0.87 - 1.01) | 0.73 (0.59 - 0.91) |
| Sleep disorder, apnea | 11248 (33.5) | 1297 (53.1) | 2030 (78.8) | 308 (107.9) | 1.54 (1.45 - 1.63) | 1.65 (1.57 - 1.74) | 2.21 (1.97 - 2.48) |
| **Musculoskeletal** |  |  |  |  |  |  |  |
| Joint pain | 27337 (86.3) | 2666 (116.9) | 4026 (149.9) | 565 (189.1) | 1.34 (1.29 - 1.39) | 1.26 (1.21 - 1.30) | 1.59 (1.46 - 1.73) |
| Dorsalgia (back pain) | 24899 (80.0) | 2528 (115.0) | 3586 (134.1) | 480 (164.6) | 1.40 (1.35 - 1.46) | 1.24 (1.20 - 1.29) | 1.50 (1.37 - 1.65) |
| Myalgia | 4355 (12.3) | 483 (18.9) | 692 (20.1) | 105 (26.9) | 1.45 (1.32 - 1.60) | 1.23 (1.13 - 1.34) | 1.56 (1.28 - 1.90) |
| **General / Other** |  |  |  |  |  |  |  |
| Fatigue | 15079 (44.6) | 1899 (82.4) | 2184 (70.4) | 367 (112.6) | 1.74 (1.66 - 1.82) | 1.15 (1.09 - 1.20) | 1.76 (1.59 - 1.96) |
| Liver abnormality | 4991 (14.1) | 529 (20.0) | 1153 (34.6) | 151 (39.4) | 1.35 (1.24 - 1.48) | 1.55 (1.45 - 1.66) | 1.71 (1.45 - 2.01) |
| Kidney injury | 3796 (10.8) | 276 (10.5) | 790 (23.9) | 85 (22.4) | 1.12 (0.99 - 1.26) | 1.21 (1.11 - 1.31) | 1.30 (1.05 - 1.62) |
| Atopic dermatitis | 2280 (6.4) | 186 (6.9) | 264 (7.5) | 23 (5.5) | 1.02 (0.88 - 1.19) | 0.96 (0.84 - 1.10) | 0.67 (0.44 - 1.02) |

CI: confidence interval. Baseline obesity was assessed as having a diagnosis between January 1, 2019 and index date. Models were adjusted for age (18-24, 25-34, 35-44, 45-54, 55-64, 65+), sex (male, female), region (northeast, north central, south, west), diagnosis time (January-March 2020, April-June 2020, July-September 2020, October-December 2020, January-March 2021, April-July 2021), and comorbidities (congestive heart failure, cardiac arrhythmias, valvular disease, pulmonary circulation disorders, peripheral vascular disorders, paralysis, other neurological disorders, chronic pulmonary disease, hypothyroidism, renal failure, liver disease, peptic ulcer disease, HIV/AIDS, lymphoma, metastatic cancer, solid tumor without metastasis, rheumatoid arthritis, coagulopathy, weight loss, fluid and electrolyte disorder, blood loss anemia, deficiency anemia, alcohol abuse, drug abuse, psychoses, depression, myocardial infraction, cerebrovascular disease, dementia, hemiplegia/paraplegia).

**Supplementary Table 7.** Rates and hazard ratios for post-acute sequelae of COVID-19 by baseline metabolic condition and COVID-19 infection, PSM matched cohorts.

| Outcomes | # of cases (Incidence per 1,000 person-years) |  |  |  | Adjusted Hazard Ratio (95% CI) |  |  |
| --- | --- | --- | --- | --- | --- | --- | --- |
|  | None | COVID-19 | Any metabolic | Both | COVID-19 vs None | Any metabolic vs None | Both vs None |
| **Cardiovascular** |  |  |  |  |  |  |  |
| Heart failure | 1592 (5.5) | 111 (5.5) | 2994 (30.2) | 250 (25.8) | 1.39 (1.04 - 1.85) | 2.14 (1.99 - 2.29) | 1.81 (1.47 - 2.22) |
| Myocardial infraction | 538 (1.9) | 41 (2.0) | 991 (9.3) | 99 (9.3) | 1.19 (0.76 - 1.87) | 1.74 (1.53 - 1.97) | 1.74 (1.25 - 2.44) |
| Stroke | 400 (1.4) | 26 (1.3) | 712 (6.6) | 79 (7.4) | 1.10 (0.63 - 1.91) | 1.57 (1.36 - 1.82) | 1.91 (1.30 - 2.82) |
| Chest pain | 7735 (27.7) | 968 (51.9) | 6045 (62.8) | 836 (95.2) | 1.78 (1.60 - 1.97) | 1.48 (1.42 - 1.55) | 2.06 (1.84 - 2.31) |
| Deep vein thrombosis | 587 (2.0) | 56 (2.7) | 649 (6.0) | 104 (9.7) | 1.32 (0.89 - 1.96) | 1.24 (1.09 - 1.42) | 1.65 (1.20 - 2.27) |
| Tachycardia | 2258 (7.9) | 334 (16.6) | 1907 (18.2) | 296 (28.9) | 2.25 (1.85 - 2.72) | 1.50 (1.39 - 1.63) | 1.96 (1.61 - 2.39) |
| Arrhythmia | 1534 (5.4) | 166 (8.1) | 1859 (18.4) | 191 (18.8) | 1.61 (1.26 - 2.06) | 1.25 (1.16 - 1.36) | 1.36 (1.09 - 1.69) |
| Angina pectoris | 386 (1.3) | 44 (2.1) | 722 (6.7) | 88 (8.2) | 1.53 (0.96 - 2.45) | 2.00 (1.74 - 2.32) | 1.93 (1.34 - 2.79) |
| **Pulmonary** |  |  |  |  |  |  |  |
| Respiratory failure | 606 (2.1) | 104 (5.2) | 1105 (10.4) | 171 (17.5) | 3.38 (2.27 - 5.02) | 1.50 (1.33 - 1.69) | 2.07 (1.60 - 2.69) |
| Pulmonary embolism | 259 (0.9) | 27 (1.3) | 363 (3.4) | 65 (6.1) | 1.19 (0.68 - 2.08) | 1.62 (1.33 - 1.97) | 2.96 (1.81 - 4.84) |
| Dyspnea | 5650 (20.2) | 814 (46.3) | 5688 (58.8) | 768 (100.7) | 2.41 (2.14 - 2.72) | 1.59 (1.52 - 1.66) | 2.45 (2.18 - 2.77) |
| **Neurological** |  |  |  |  |  |  |  |
| Nerve/nerve root/plexus disorder | 3005 (10.6) | 311 (15.4) | 2524 (24.2) | 337 (32.6) | 1.42 (1.19 - 1.68) | 1.47 (1.37 - 1.56) | 1.75 (1.46 - 2.08) |
| Anosmia (loss taste/smell) | 749 (2.6) | 123 (6.2) | 296 (2.7) | 69 (6.5) | 2.47 (1.79 - 3.42) | 1.12 (0.95 - 1.33) | 2.26 (1.48 - 3.46) |
| Other neurological | 156 (12.8) | 22 (10.4) | 198 (25.5) | 55 (36.9) | 0.83 (0.40 - 1.71) | 0.99 (0.75 - 1.29) | 1.52 (0.85 - 2.74) |
| Amnesia (loss of memories) | 1910 (6.7) | 207 (10.2) | 1949 (18.4) | 257 (24.7) | 1.59 (1.28 - 1.98) | 1.29 (1.19 - 1.40) | 1.56 (1.28 - 1.90) |
| **Menta behavioral** |  |  |  |  |  |  |  |
| Anxiety disorder | 16474 (65.7) | 1574 (93.5) | 6609 (76.3) | 840 (103.4) | 1.38 (1.28 - 1.49) | 1.17 (1.13 - 1.22) | 1.35 (1.22 - 1.50) |
| Mood disorder | 9168 (34.4) | 867 (46.7) | 4529 (48.7) | 547 (60.7) | 1.36 (1.23 - 1.51) | 1.35 (1.29 - 1.42) | 1.54 (1.35 - 1.76) |
| Psychotic disorder | 235 (0.8) | 20 (1.0) | 146 (1.3) | 24 (2.2) | 2.01 (0.94 - 4.30) | 1.54 (1.14 - 2.08) | 4.50 (1.71 - 11.82) |
| Substance misuse | 5567 (19.9) | 417 (21.2) | 2870 (28.7) | 230 (22.6) | 1.18 (1.02 - 1.35) | 1.31 (1.24 - 1.39) | 1.05 (0.87 - 1.26) |
| Sleep disorder, apnea | 7599 (27.7) | 809 (42.3) | 5679 (65.3) | 796 (97.4) | 1.53 (1.37 - 1.71) | 1.56 (1.49 - 1.62) | 2.05 (1.83 - 2.30) |
| **Musculoskeletal** |  |  |  |  |  |  |  |
| Joint pain | 19260 (74.4) | 1825 (102.7) | 12103 (143.3) | 1406 (175.2) | 1.43 (1.33 - 1.53) | 1.31 (1.27 - 1.34) | 1.51 (1.39 - 1.63) |
| Dorsalgia (back pain) | 18184 (71.6) | 1790 (104.8) | 10301 (122.9) | 1218 (155.7) | 1.42 (1.32 - 1.52) | 1.28 (1.24 - 1.32) | 1.50 (1.38 - 1.64) |
| Myalgia | 3258 (11.5) | 350 (18.1) | 1789 (16.9) | 238 (23.4) | 1.60 (1.35 - 1.89) | 1.16 (1.08 - 1.25) | 1.50 (1.23 - 1.84) |
| **General / Other** |  |  |  |  |  |  |  |
| Fatigue | 11067 (40.6) | 1338 (75.9) | 6196 (64.2) | 928 (106.9) | 1.67 (1.54 - 1.82) | 1.17 (1.12 - 1.21) | 1.66 (1.50 - 1.84) |
| Liver abnormality | 2987 (10.5) | 311 (15.4) | 3157 (30.7) | 369 (36.6) | 1.60 (1.34 - 1.92) | 1.84 (1.73 - 1.96) | 1.91 (1.60 - 2.28) |
| Kidney injury | 1592 (5.5) | 111 (5.5) | 2994 (30.2) | 250 (25.8) | 1.39 (1.04 - 1.85) | 2.14 (1.99 - 2.29) | 1.81 (1.47 - 2.22) |
| Atopic dermatitis | 1735 (6.1) | 157 (7.7) | 809 (7.5) | 52 (4.8) | 1.31 (1.04 - 1.67) | 1.09 (0.98 - 1.21) | 0.70 (0.49 - 1.01) |

PSM: propensity score matching. CI: confidence interval.

**Supplementary Table 8.**
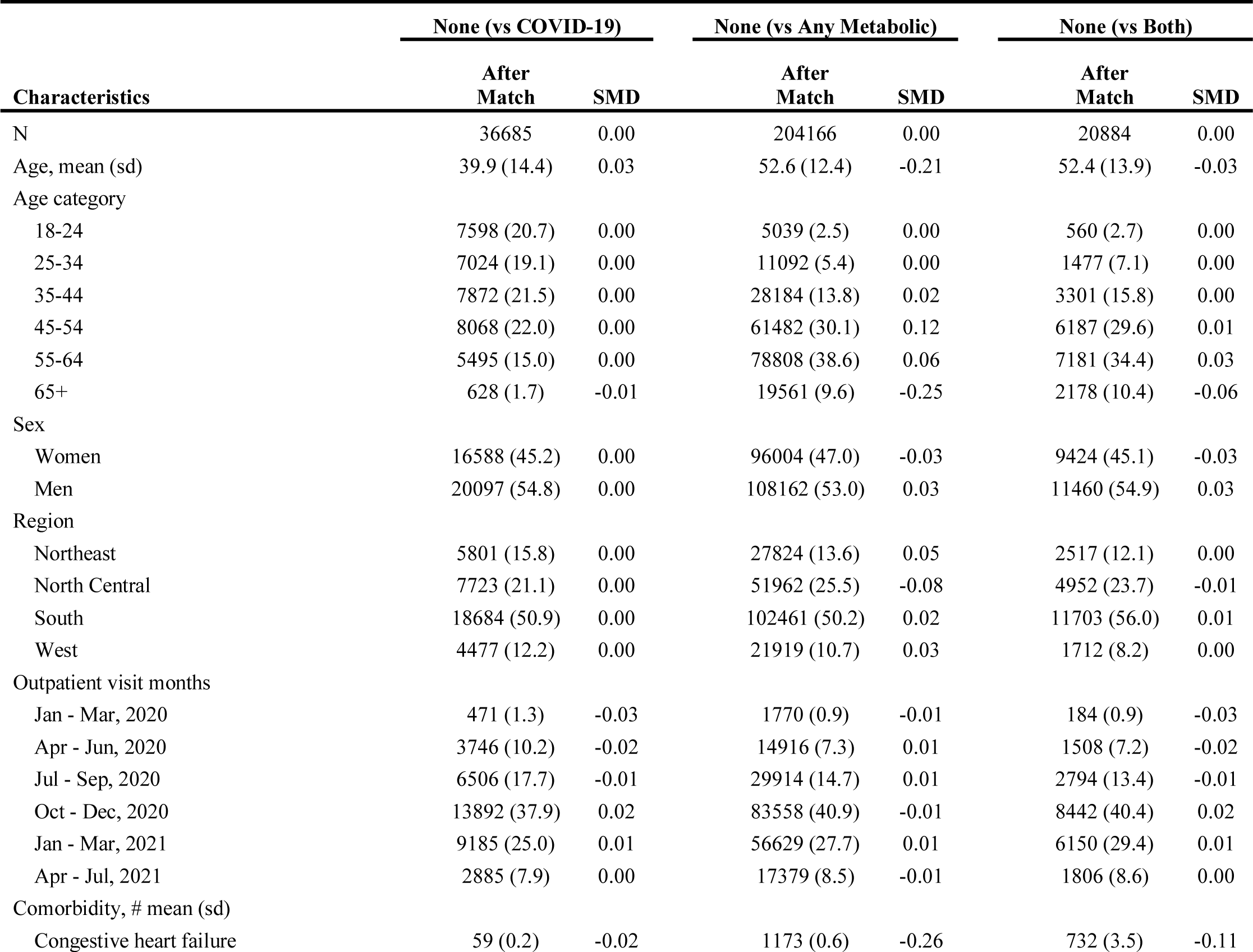
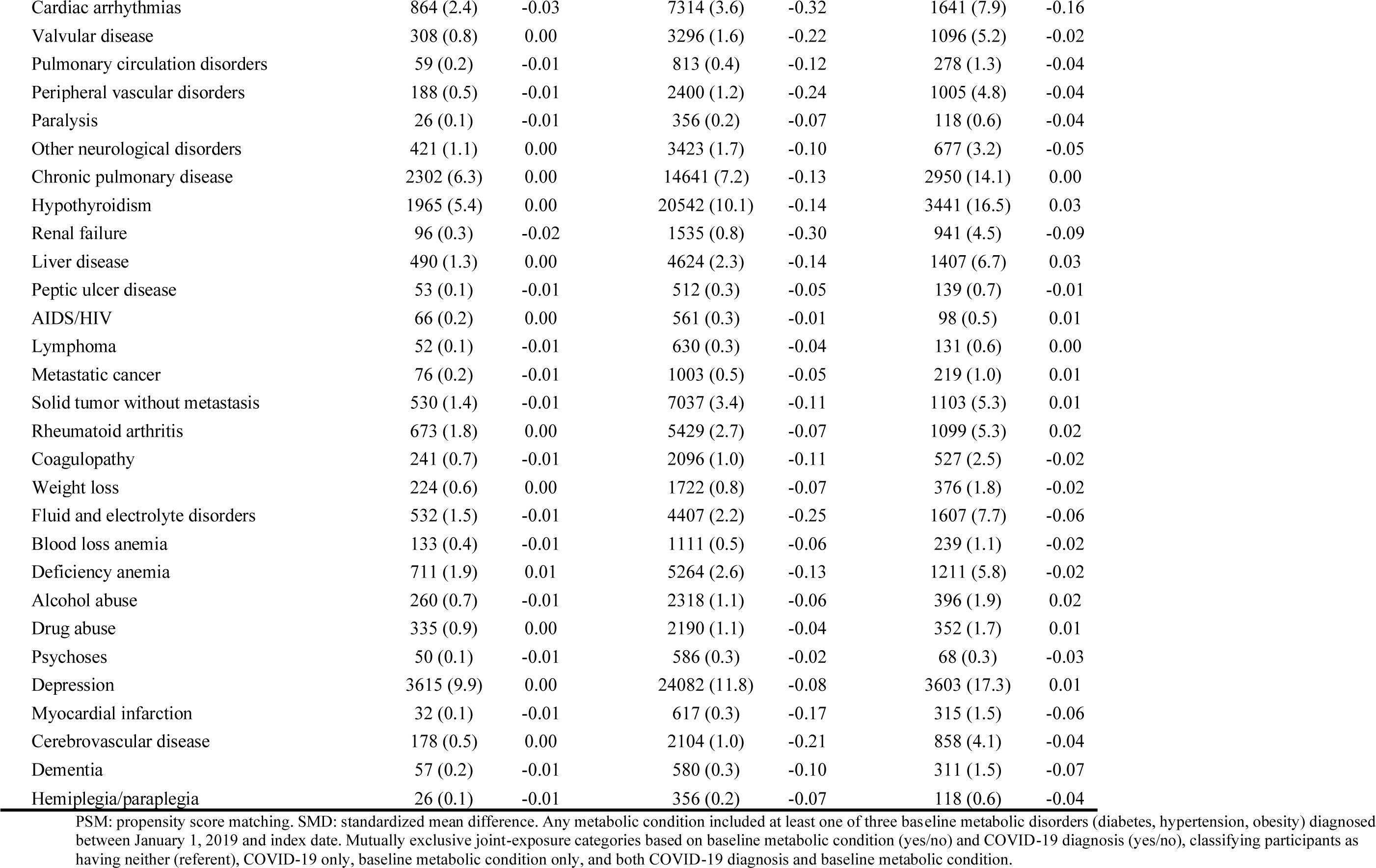
PSM matched reference sample characteristics by COVID-19 infection and baseline metabolic condition.

## Notes

### Author Declarations

Ethical approval was granted by the Boston University/Boston Medical Center Institutional Review Board.

